# Toward Reliable Thalamic Segmentation: a rigorous evaluation of automated methods for structural MRI

**DOI:** 10.1101/2025.09.09.25335439

**Authors:** Georgios P. D. Argyropoulos, Christopher R. Butler, Manojkumar Saranathan

**Affiliations:** Memory Research Group, Nuffield Department of Clinical Neurosciences, University of Oxford, Oxford, United Kingdom; Division of Psychology, Faculty of Natural Sciences, University of Stirling, Scotland, United Kingdom; Department of Brain Sciences, Imperial College London, London W12 0NN, UK; Departamento de Neurología, Pontificia Universidad Católica de Chile, Avda. Libertador Bernando O’Higgins 340, Santiago, Chile; Department of Radiology, University of Massachusetts Chan Medical School, Worcester, MA, USA

**Keywords:** Thalamus, segmentation, multi-atlas segmentation, thalamic nuclei, THOMAS, HIPS- THOMAS, FreeSurfer, MRI, DTI

## Abstract

Automated thalamic nuclear segmentation has contributed towards a shift in neuroimaging analyses from treating the thalamus as a homogeneous, passive relay, to a set of individual nuclei, embedded within distinct brain-wide circuits. However, many studies continue to widely rely on FreeSurfer’s segmentation of T1-weighted structural MRIs, despite their poor intrathalamic nuclear contrast. Meanwhile, a convolutional neural network tool has been developed for FreeSurfer, using information from both diffusion and T1-weighted MRIs. Another popular thalamic nuclear segmentation technique is HIPS-THOMAS, a multi-atlas-based method that leverages white-matter-like contrast synthesized from T1-weighted MRIs. However, rigorous comparisons amongst methods remain scant, and the thalamic atlases against which these methods have been assessed have their own limitations. These issues may compromise the quality of cross-species comparisons, structural and functional connectivity studies in health and disease, as well as the efficacy of neuromodulatory interventions targeting the thalamus. Here, we report, for the first time, comparisons amongst HIPS-THOMAS, the standard FreeSurfer segmentation, and its more recent development, against two thalamic atlases as silver-standard ground-truths. We used two cohorts of healthy adults, and one cohort of patients in the chronic phase of autoimmune limbic encephalitis. In healthy adults, HIPS-THOMAS surpassed, not only the standard FreeSurfer segmentation, but also its more recent, diffusion-based update. The improvements made with the latter relative to the former were limited to a few nuclei. Finally, the standard FreeSurfer method underperformed, relative to the other two, in distinguishing between patients and healthy controls based on the affected anteroventral and pulvinar nuclei. In light of the above findings, we provide recommendations on the use of automated segmentation methods of the human thalamus using structural brain imaging.

## Introduction

There is growing recognition of the need for neuroimaging studies to stop treating the thalamus as a single, homogeneous, passive relay, and instead examine its nuclei as separate, highly specialised structures supporting multiple cognitive processes (Segobin et al., 2024). Until recently, distinguishing human thalamic nuclei was only feasible via post-mortem histology or manual delineation of ex vivo MRIs. While the segmentation of all nuclei (n>60) remains beyond the capacity afforded by current MRI resolution, developments since the second half of the last decade have enabled the automated segmentation of several nuclei from diffusion, functional, and structural MRI data (Battistella et al., 2017; Iglesias et al., 2018; Su et al., 2019; Tregidgo et al., 2023; Umapathy et al., 2022; van Oort et al., 2018; Vidal et al., 2024).

The most widespread of these methods is the Bayesian-based segmentation of T1w-MRI incorporated into FreeSurfer’s segmentation (FS-T1), which is based on probabilistic maps of thalamic nuclei determined from ex vivo MRIs and post-mortem histology samples (Iglesias et al., 2018). It is used, not only for volumetry (Nakhid et al., 2023; Trufanov et al., 2021; Mørch-Johnsen et al., 2023; Lee, Seo, Lee, et al., 2020; Shin et al., 2019; Xiong et al., 2021), but also for ROI definition in task-based fMRI analyses (Gentile et al., 2024), and functional (Velioglu et al., 2023) and structural connectivity analyses (Liu et al., 2022). Another popular method, especially in the neurosurgical world, is THalamus Optimized Multi-Atlas Segmentation method (Su et al., 2019) or THOMAS. This leverages the improved intrathalamic contrast of white-matter nulled (WMn) T1-w MRI sequences (Su et al., 2019) like FGATIR. A recent variant, HIPS-THOMAS (Vidal et al., 2024), uses HIstogram-based Polynomial Synthesis (HIPS), which synthesizes WMn image contrast from standard T1w-MRIs, removing the need for specialized sequences. WMn-MRI has been shown to generate more accurate segmentation (Umapathy et al., 2022) relative to T1w-MRIs.

A more recent development has been the convolutional neural network tool for FreeSurfer (Tregidgo et al., 2023) (FS-DTI), which uses information from both diffusion and T1w-MRI (we refer to this as FS-DTI). Given its implementation in FreeSurfer and the improved visualization of ventral and medial thalamic boundaries, we expect this method to be more accurate and become rapidly adopted by the community – although reports of its usage for thalamic nuclear segmentation remain very limited to date.

Despite these advances, there is no consensus on the thalamic nuclei that can be reliably segmented or the techniques that should be leveraged to enhance their delineation (Segobin et al., 2024). While validation analyses of FS-T1 have showed poor results (Su et al., 2019; Williams et al., 2022) and lower quality relative to HIPS-THOMAS for several nuclei (Williams et al., 2024), comparisons between approaches are scant (Iglehart et al., 2020; Williams et al., 2024). Importantly, the two state-of-the-art structural-based methods-HIPS-THOMAS and FS-T1-have not been compared with the recent FS-DTI improvement. Moreover, the multiplicity of methods available and the lack of objective performance measures leave room for cherry-picking (Segobin et al., 2024). Comparative neuroanatomical studies, robust replication of findings in cognitive/clinical neuroscience, as well as favourable treatment outcomes of neuromodulatory interventions targeting the thalamus are all dependent on the quality of thalamic segmentation methods.

Gold-standard manual segmentations (e.g., Su et al., 2019) are not an easily attainable ground-truth when comparing automated segmentation methods on large datasets. For this reason, the Krauth-Morel atlas (K-M) (Krauth et al., 2010), a digital representation of the Morel stereotactic atlas (Morel, 2007; Morel et al., 1997) brought to MNI152 space (Jakab et al., 2012), is often used as a “silver-standard” reference (Williams et al., 2022, 2024). However, an obvious disadvantage is that the Morel atlas has been constructed on a limited series of post-mortem data (9 thalamic blocks from 5 normal human brains), thus presenting with very little variability in anatomy. More relevantly, recent work has highlighted limitations in the quality of the MNI-space K-M atlas, especially for segmentations interfacing with large white matter bundles or cerebrospinal fluid (Brun et al., 2022). This calls into question the utility of the K-M atlas as a silver-standard ground truth in comparing segmentation methods (Williams et al., 2024). Brun et al. (2022) also proposed a new 7TAMIbrainDGN atlas, which contains 24 deep grey matter (including 12 thalamic) nuclei per hemisphere on their high-contrast/resolution T1w 7TAMIbrainT1w_30 template.

In this work, we conducted rigorous performance comparisons of FS-T1, FS-DTI, and HIPS-THOMAS in two separate cohorts of healthy adults with T1w-MRI and diffusion MRI data. To address the issue of atlas variability, they were evaluated against two thalamic atlases: the MNI-space K-M atlas, and an MNI-space version that we created for the 7TAMIbrainDGN atlas. Lastly, we also compared FS-T1, FS-DTI, and HIPS-THOMAS on their efficacy in distinguishing between healthy adults and patients in the chronic phase of autoimmune limbic encephalitis (aLE) - a non-neurogenerative condition that is mainly associated with inflammation and subsequent atrophy in medial temporal lobe structures, primarily the hippocampus (Argyropoulos et al., 2019).

## Methods

### Datasets

#### MAP35

MAP35 comprised 35 healthy adults (age: mean=55.14; SD=14.04 years; 25M:10F), recruited in the Memory and Amnesia Project (Nuffield Department of Clinical Neurosciences, University of Oxford) (Argyropoulos et al., 2025). T1w-MRIs were acquired using a MPRAGE sequence (echo time = 4.7 ms, repetition time = 2040 ms, 8° flip angle, field of view = 192 mm, voxel size = 1×1×1 mm). Diffusion MRI involved a single-shot EPI sequence (64 slices; slice thickness = 2 mm, 0 gap; axially acquired; 64 directions at b = 1,500 s/mm^2^, repetition time = 8,900 ms; echo time = 94.8 ms; voxel size = 2 × 2 × 2 mm; field of view = 192 × 192 mm), along with a no-diffusion-weighted image (b = 0 s/mm^2^).

#### HCP35

We sought to replicate our findings with a publicly available MRI dataset of equivalent size – for this reason, we selected the “MGH HCP Adult Diffusion” dataset (henceforth “HCP35”) of the Human Connectome Project. HCP35 comprises diffusion and structural imaging data from 35 healthy young adults (age: range=20-59; estimated mean=31.57; estimated SD=8.52; 16F:19M). Details on T1w-MRI and Diffusion MRI scans and preprocessing can be found here: https://humanconnectome.org/study/hcp-young-adult/document/mgh-adult-diffusion-data-acquisition-details/. Of those, however, we excluded 2 subjects’ datasets (mgh_1035, as this dataset lacked preprocessed data, and mgh_1020, as the FS-DTI segmentation failed upon inspection, also reflected in the total thalamic volumes estimated by each method). This is shown in Supplementary Table 1.

**Table 1:** Binomial logistic regression analyses, one per method and segmentation; **key:** p-corr(18): the p-value was adjusted to correct for the number of multiple comparisons made (n=18), using the Holm-Bonferroni sequential correction method (Holm, 1979); AV: anteroventral nucleus; MD-Pf: mediodorsal-parafascicular nucleus; Pul: pulvinar nucleus; L/R: Left, Right hemisphere; FS-T1: T_1_-based FreeSurfer segmentation (Iglesias et al., 2018); FS-DTI: FreeSurfer’s joint segmentation of thalamic nuclei from T1 scan and DTI (Tregidgo et al., 2023); HIPS-THOMAS: Thalamus Optimized Multi-atlas Segmentation using Histogram-based Polynomial Synthesis (Vidal et al., 2024); Seg.: Segmentation; Hem.: Hemisphere; SE: Standard Error; AUC: Area Under the Curve.

| Seg. | Hem. | Method | AUC | Coefficient | SE | z | p-corr(18) |
| --- | --- | --- | --- | --- | --- | --- | --- |
| AV | L | HIPS-THOMAS | 0.80 | 1.28 | 0.35 | 3.71 | 0.003 |
|  |  | FS-DTI | 0.82 | 1.68 | 0.42 | 4.01 | 0.001 |
|  |  | FS-T1 | 0.62 | 0.28 | 0.25 | 1.11 | 0.685 |
|  | R | HIPS-THOMAS | 0.83 | 1.81 | 0.45 | 4.00 | 0.001 |
|  |  | FS-DTI | 0.82 | 1.74 | 0.45 | 3.85 | 0.002 |
|  |  | FS-T1 | 0.57 | 0.22 | 0.24 | 0.89 | 0.685 |
| MD-Pf | L | HIPS-THOMAS | 0.79 | 1.27 | 0.35 | 3.63 | 0.004 |
|  |  | FS-DTI | 0.77 | 1.26 | 0.37 | 3.40 | 0.007 |
|  |  | FS-T1 | 0.71 | 0.82 | 0.28 | 2.90 | 0.019 |
|  | R | HIPS-THOMAS | 0.76 | 1.15 | 0.35 | 3.28 | 0.008 |
|  |  | FS-DTI | 0.78 | 1.37 | 0.39 | 3.48 | 0.006 |
|  |  | FS-T1 | 0.76 | 1.16 | 0.34 | 3.43 | 0.007 |
| Pul | L | HIPS-THOMAS | 0.80 | 1.28 | 0.35 | 3.67 | 0.003 |
|  |  | FS-DTI | 0.76 | 1.10 | 0.33 | 3.37 | 0.007 |
|  |  | FS-T1 | 0.59 | 0.29 | 0.24 | 1.21 | 0.685 |
|  | R | HIPS-THOMAS | 0.76 | 1.26 | 0.38 | 3.30 | 0.008 |
|  |  | FS-DTI | 0.73 | 1.13 | 0.37 | 3.05 | 0.014 |
|  |  | FS-T1 | 0.61 | 0.36 | 0.25 | 1.44 | 0.599 |

#### Patients

For our Receiver Operating Characteristic (ROC) analyses (*infra*), we also used the data of the 37/38 patients (scanned using the protocol of the MAP35 dataset) reported in Argyropoulos et al. (2025) (26M:11F; age at imaging: mean = 61.14; SD = 14.01 years; controls vs. patients: M:F ratio: χ^2^ = 0.01, p=0.914; age at imaging: t(70) = 1.81, p = 0.074), who had undergone both T1w-MRI and diffusion MRI. These patients were in the chronic phase of aLE, which is primarily characterised by hippocampal atrophy and residual episodic memory impairment (Argyropoulos et al., 2019).

### Native-space segmentation

#### FS-T1

We used the output of FreeSurfer’s (v. 7.4.1-20230614-7eb8460) “recon-all” (T1w-MRI parcellation; command: “recon-all -i T1.nii.gz -s T1 -all”) to segment the thalamic nuclei with the algorithm (“segmentThalamicNuclei.sh”; command: “segment_subregions thalamus -- cross T1”) described in Iglesias et al. (2018). Thalamic segmentations were converted from FS to native space (“mri_label2vol”), and from “.mgz” to “.nii” format (“mri_convert”). The segmented nuclei comprise the anteroventral (AV), the centrolateral (CL), centromedian (CM), central medial (CeM), lateral geniculate (LGN), lateral posterior (LP), laterodorsal (LD), limitans-suprageniculate (LSg), medial geniculate (MGN), mediodorsal lateral-parvocellular (MDl), mediodorsal medial-magnocellular (MDm), reuniens-medial ventral (MVRe), paracentral (Pc), parafascicular (Pf), paratenial (Pt), pulvinar anterior (PuA), pulvinar inferior (PuI), pulvinar lateral (PuL), pulvinar medial (PuM), ventral anterior (VA), ventral anterior magnocellular (VAmc), ventral lateral anterior (VLa), ventral lateral posterior (VLp), ventromedial (VM), ventral posterolateral nuclei (VPL).

#### FS-DTI

For FS-DTI (Tregidgo et al., 2023), we provided the following inputs to the command (“mri_segment_thalamic_nuclei_dti_cnn --t1 norm.mgz --aseg aseg.mgz --fa dtifit_FA.nii.gz - -v1 dtifit_V1.nii.gz --o thal_cnn_outputSegmentation.nii.gz --vol thal_cnn_volumes.csv -- model /usr/local/freesurfer/7.4.1/models/thalseg_1.1.h5 --threads 3”): a bias-corrected, whole-brain structural T1w-MRI (“norm.mgz”), a whole-brain segmentation (“aseg.mgz”-output from “recon-all”, *supra*), a fractional anisotropy volume (“dtifit_FA.nii.gz”) and a 4D volume containing the principal direction vector for each DTI voxel (“dtifit_V1.nii.gz”). The last two were derived with TRACULA (TRActs Constrained by UnderLying Anatomy; Maffei et al., 2021; Yendiki et al., 2011) for MAP35, and with DTIFit in FSL (v. 6.0.7.8) for HCP35, as the data from the latter dataset were already preprocessed with gradient non-linearity correction, motion correction, eddy-current correction, b-vector correction (https://humanconnectome.org/study/hcp-young-adult/document/mgh-adult-diffusion-data-acquisition-details/; note that TRACULA also calls DTIFit: https://surfer.nmr.mgh.harvard.edu/fswiki/FsTutorial/TraculaOutputs). The algorithm also used the FS-T1-based segmentation. Each subject’s norm.mgz was registered, without resampling, to diffusion space. The segmented nuclei were the same as those of FS-T1, with the following exceptions: two segmentations are generated for PuM: a medial segment, PuMm, and a lateral one (PuMl); Pc, VM and Pt segmentations are not generated, as they do not appear in all of the training examples for the FS-DTI’s CNN, so these are excluded to improve the robustness of the network (https://surfer.nmr.mgh.harvard.edu/fswiki/ThalamicNucleiDTI).

#### HIPS-THOMAS

We used HIPS-THOMAS v.2 (command: “docker run -it --rm --name sthomas -v ${PWD}:/data -w /data --user ${UID}:$(id -g) anagrammarian/sthomastest hipsthomas.sh -v -t1 -i T1.nii.gz”) to segment the thalamus. The segmented nuclei comprise the AV, VA, VLa, VLp, VPL, Pul, LGN, MGN, CM, MD-Pf, and habenular (Hb) nuclei as well as CL.

#### Segmentation preparation

To enable direct comparisons amongst the three methods, FS segmentations and K-M ROIs were combined to match the nomenclature employed in HIPS-THOMAS. Segmentations not generated by HIPS-THOMAS (e.g., LD) or FS-T1/FS-DTI (e.g., Hb) were omitted. This resulted in 22 segmentations (11/hemisphere) (Supplementary Table 2). We followed the methodology described in Williams et al. (2024), with the only exception being the addition of centrolateral (CL) nucleus in the new HIPS-THOMAS version. We repeated these comparisons using the 7TAMIbrainDGN atlas as a second silver-standard reference. As the 7TAMIbrainDGN is in its own reference space, the thalamic nuclei of interest from this atlas were warped, as above, to FSL’s MNI152_T1_1mm_brain for the purposes of comparing the three segmentation methods (Supplementary Table 3). For brevity, we henceforth refer to this custom-made MNI version of the 7TAMIbrainDGN as “7TAMI”.

**Table 2:**
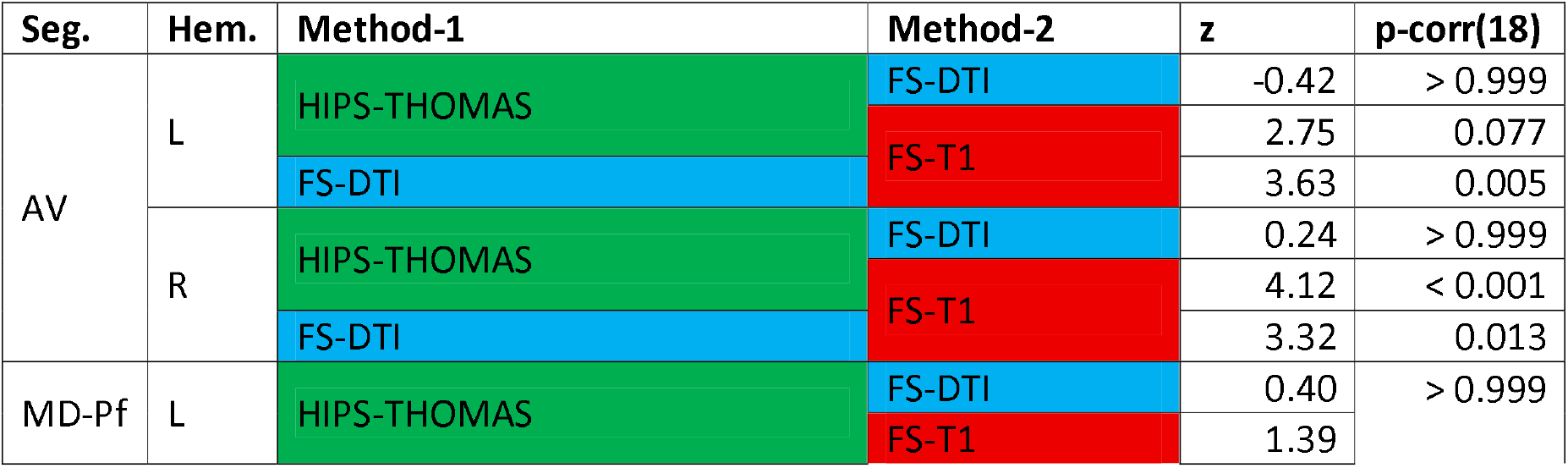
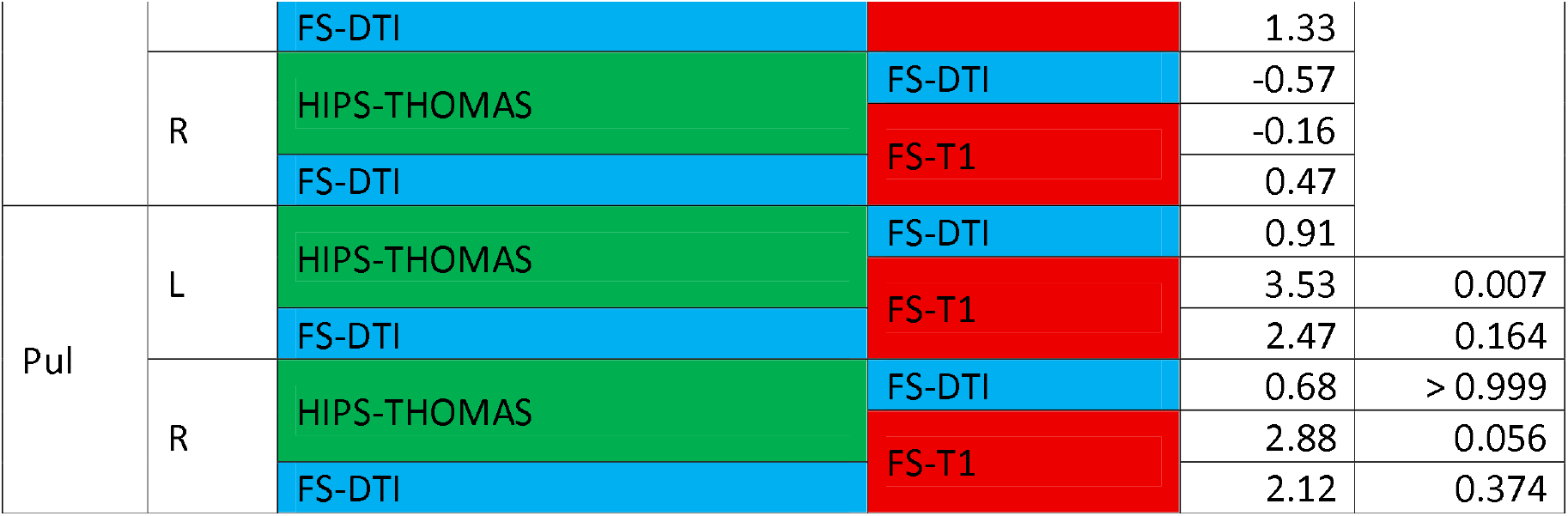
DeLong comparisons of segmentation methods on AUCs for L/R AV, MD-Pf, and Pul; **key:** p-corr(18): the p-value was adjusted to correct for the number of multiple comparisons made (n=18), using the Holm-Bonferroni sequential correction method (Holm, 1979); AV: anteroventral segmentation; MD-Pf: mediodorsal-parafascicular segmentation; Pul: pulvinar segmentation; L/R: Left, Right hemisphere; FS-T1: T_1_-based FreeSurfer segmentation (Iglesias et al., 2018); FS-DTI: FreeSurfer’s joint segmentation of thalamic nuclei from T1 scan and DTI (Tregidgo et al., 2023); HIPS-THOMAS: Thalamus Optimized Multi-atlas Segmentation using Histogram-based Polynomial Synthesis (Vidal et al., 2024); Seg.: Segmentation; Hem.: Hemisphere.

Following thalamic segmentation in native space, each T1w-MRI underwent brain extraction (FSL’s BET), and an affine matrix was calculated with FSL’s FLIRT (Jenkinson et al., 2002) for rigid/affine linear registration to MNI space (initial global alignment; template: MNI152_T1_1mm_brain: https://git.fmrib.ox.ac.uk/fsl/data_standard/-/raw/master/MNI152_T1_1mm_brain.nii.gz?ref_type=heads&inline=false). Using that affine matrix, nonlinear registration was conducted for the T1w-MRI to MNI space (FSL’s FNIRT; Andersson et al., 2008), generating warps from native to MNI space and from MNI to native space. Using nearest-neighbour interpolation, the former was applied to register native-space segmentations from all three methods to MNI space, and the latter was used for all K-M ROIs in MNI space (Jakab et al., 2012; Krauth et al., 2010), separately for each subject of each dataset, bringing them to each subject’s native anatomical space. The same warps were used when comparisons were repeated with 7TAMI as the second ground-truth.

#### Statistical analysis

For both the native-space and the MNI analyses and for both reference atlases, we used two complementary dependent variables: Dice coefficients and Average Hausdorff Distances (AHDs). The Dice coefficient, an overlap-based metric, is defined as *“Dice(S,R)=2*[(IZS⍰R⍰)/(⍰S⍰+⍰R⍰)]”*, wherein *“S”* is a certain segmentation from FS-T1, FS-DTI, or HIPS-THOMAS, *“R”* is the corresponding same-space ground-truth reference K-M/7TAMI segmentation, *“S⍰R”* corresponds to the set of voxels that are shared by S and R, and *“⍰⍰”* measures the number of voxels in the segmentation. As elsewhere (Pajula et al., 2012; Williams et al., 2024), we used the following value cut-offs to discuss segmentation accuracy: Dice = 0: “no agreement”; 0 < Dice < 0.2: “slight agreement”; 0.2 ≤ Dice < 0.4: “fair agreement”; 0.4 ≤ Dice < 0.6: “moderate agreement”; 0.6 ≤ Dice < 0.8: “substantial agreement”; 0.8 ≤ Dice ≤ 1: “almost perfect agreement”. AHD, a distance-based metric, helps address the issue with overlap-based metrics disproportionately penalising smaller, relative to larger, segmentations, for errors (Taha & Hanbury, 2015; Williams et al., 2024). We defined it as *“AHD(S,R) = max(d(S,R),d(R,S))”*, where “*d*(*S,R*)” is the average minimum distance (min ⍰*s* − *r*⍰) from voxels in S to R, “*d*(*R, S*)” is the average minimum distance (min ⍰*r* − *s*⍰) from voxels in R to S, and “*max*” refers to the largest of these two values (d(R,S), d(S,R)) being selected. Similar to Williams et al. (2024), we treat segmentations with AHD≤1mm as being of sufficiently small distance from the corresponding ground-truth reference K-M/7TAMI segmentation.

We used a series of one-way repeated-measures ANOVAs (in RStudio v. 2023.06.0 Build 421 © 2009-2023 Posit Software, PBC; R v. 4.4.3 - 2025-02-28 ucrt) to compare amongst the three segmentation methods in native space, separately for each dataset (HCP35, MAP35), and DV (Dice coefficient, AHD). We used the Holm-Bonferroni sequential correction method (Holm, 1979) to correct for the number of repeated-measures ANOVAs per dataset and metric (n=22 for K-M, n=16 for 7TAMI), as well as for the post-hoc pair-wise comparisons (n=3) between segmentation methods. The Greenhouse-Geisser correction was used for the ANOVAs to adjust the degrees of freedom whenever the assumption of sphericity was violated.

### MNI-space analyses

Following the spatial transformation of all segmentations from native to MNI space, group-level probabilistic maps were generated, per segmentation of interest (n=24; Supplementary Tables 2-3), method (n=3), and dataset/group (n=2). We determined the proportion of each group that was assigned to a segmentation at each voxel position (Najdenovska et al., 2018; Williams et al., 2024), for each of the three methods. A threshold of 0.25 was used to binarize the group-level probabilistic atlas (Williams et al., 2024). Dice coefficients and AHDs were derived for each group-level segmentation in MNI space for each method and dataset relative to the corresponding K-M or 7TAMI ground-truth reference segmentation. The calculation of Dice and AHD metrics in both native and MNI space, as well as the generation of the group-level probabilistic atlases were conducted using SPM12 (v. 7771) in Matlab (v. 2023a Update 8 - 9.14.0.2891782) 64-bit).

### “Best” method per segmentation

We applied a series (n=8) of stringent criteria to identify the “best” segmentation method amongst the three examined, for each of the segmentations in question. A method was identified as “the best” if it met all 8 of the criteria – i.e., if it surpassed the other two for both Dice (higher) and AHD metrics (lower), in both native-space (statistically) and MNI space (numerically), in both the HCP35 and MAP35 datasets. Two segmentation methods were identified as “joint-best” if they both consistently surpassed the third one on all the aforementioned criteria. Of the segmentations for which no best method was found to satisfy 8/8 criteria, we identified the methods that satisfied >4/8 and ≤7/8 criteria.

### ROC analysis

We performed a series of ROC analyses for our dataset of healthy controls (n=35 from MAP35) and patients (n=37). Volumes were residualised against age, sex, and total intracranial volume (FS’s ‘eTIV’). We used binomial logistic regression to quantify the capacity of FS-T1, FS-DTI, and HIPS-THOMAS to discriminate between healthy controls and aLE patients, on the basis of the 6 volumes (L/R MD-Pf, AV, Pul) that HIPS-THOMAS and FS-DTI had both shown volume reduction in Argyropoulos et al. (2025). We first calculated area under the curve (AUC) values for each of the three segmentation methods. We first included all 6 nuclei as predictors. However, to address collinearity issues and identify the performance of each method on each of these segmentations, we then repeated these analyses separately per method and segmentation (n=18). We also used DeLong’s tests to compare amongst the AUCs of the three methods on each of those segmentations. P-values were corrected as above.

## Results

### Native-space analyses

When using the K-M as ground truth, HIPS-THOMAS outperformed both FS-T1 and FS-DT1 for R AV, L/R CL, L/R LGN, L/R Pul, L/R VLp, and L/R VPL consistently across datasets (HCP35, MAP35) and metrics (Dice coefficients: Figure 1a, Supplementary Figure 1a, Supplementary Table 4; AHDs: Figure 2a, Supplementary Figure 2a, Supplementary Table 6). FS-DTI consistently outperformed FS-T1 and HIPS-THOMAS only on L/R MD-Pf. FS-T1 did not consistently outperform the other two for any segmentation. The above findings were conceptually replicated in a strong fashion when using the 7TAMI as ground truth (Figures 1b,2b; Supplementary Figures 1b,2b; Supplementary Tables 5, 7).

**Figure 1:**
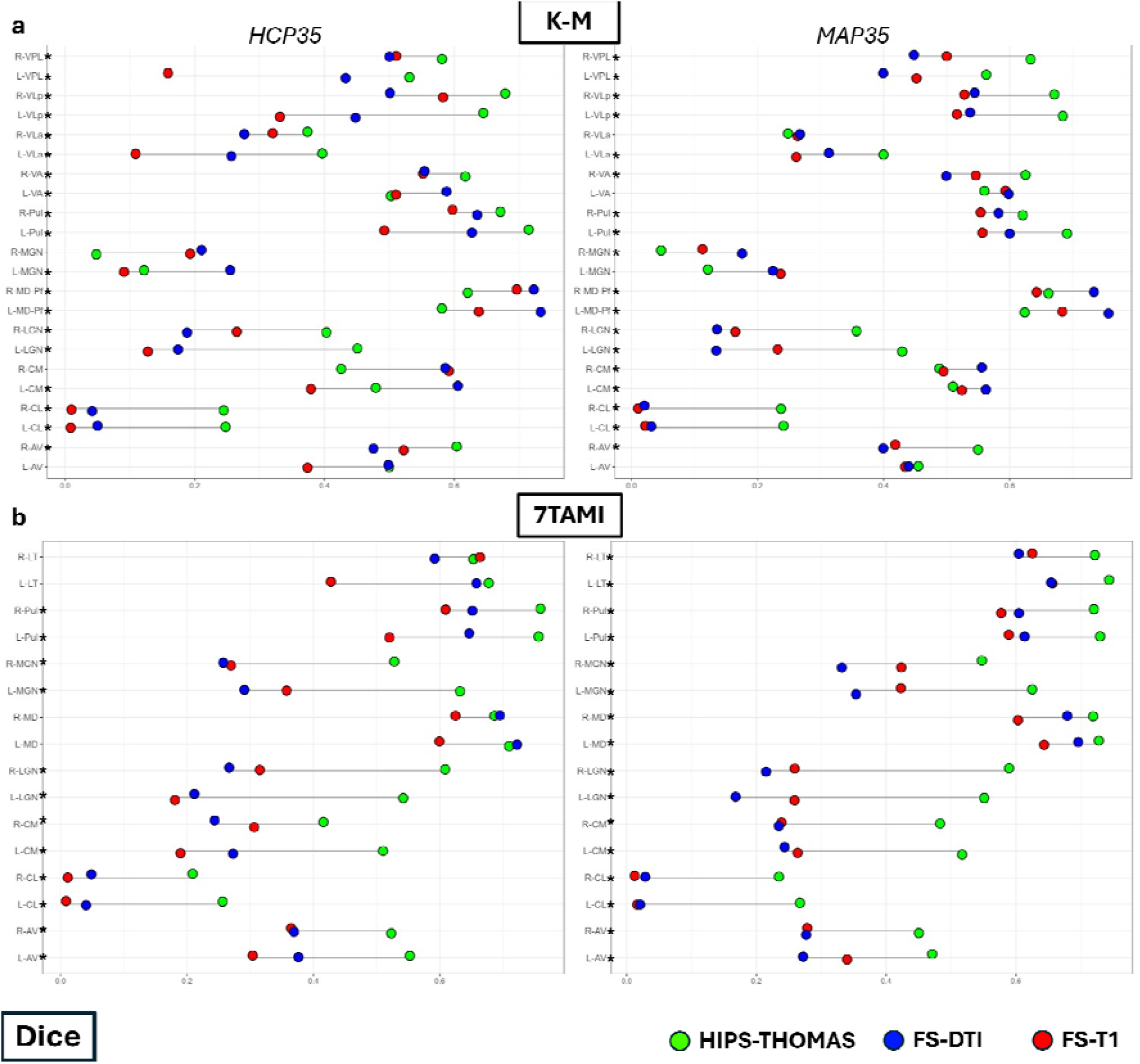
Dumbbell plots of Dice coefficients for nuclei segmented with FS-T1, FS-DTI, and HIPS-THOMAS relative to a) K-M and b) 7TAMI ground-truth reference segmentations (native space; see violin plots in Supplementary Figure 1); **key:** *: significantly higher Dice coefficients for one method relative to the other two; AV: anteroventral nucleus; LT: lateral nuclei (7TAMI); VA: ventral anterior nucleus; VLa: Ventrolateral anterior nucleus; VLp: Ventrolateral posterior nucleus; MD-Pf: mediodorsal-parafascicular nuclei; Pul: pulvinar nucleus; VPL: Ventral Posterolateral nucleus; CL: Centrolateral nucleus; CM: Centromedian nucleus; LGN: Lateral Geniculate Nucleus; MGN: Medial Geniculate Nucleus; L/R: Left, Right hemisphere; FS-T1: T_1_-based FreeSurfer segmentation (Iglesias et al., 2018); FS-DTI: FreeSurfer’s joint segmentation of thalamic nuclei from T1 scan and DTI (Tregidgo et al., 2023); HIPS-THOMAS: Thalamus Optimized Multi-atlas Segmentation using Histogram-based Polynomial Synthesis (Vidal et al., 2024); MAP35, HCP35: datasets with T1w-MRIs and diffusion MRI available; black diamond: mean; black horizontal line: median; K-M: Krauth-Morel atlas (Krauth et al., 2010); 7TAMI: the custom-made MNI version of the atlas of deep grey matter nuclei, part of the 7TAMIbrain dataset (Brun et al., 2022).

**Figure 2:**
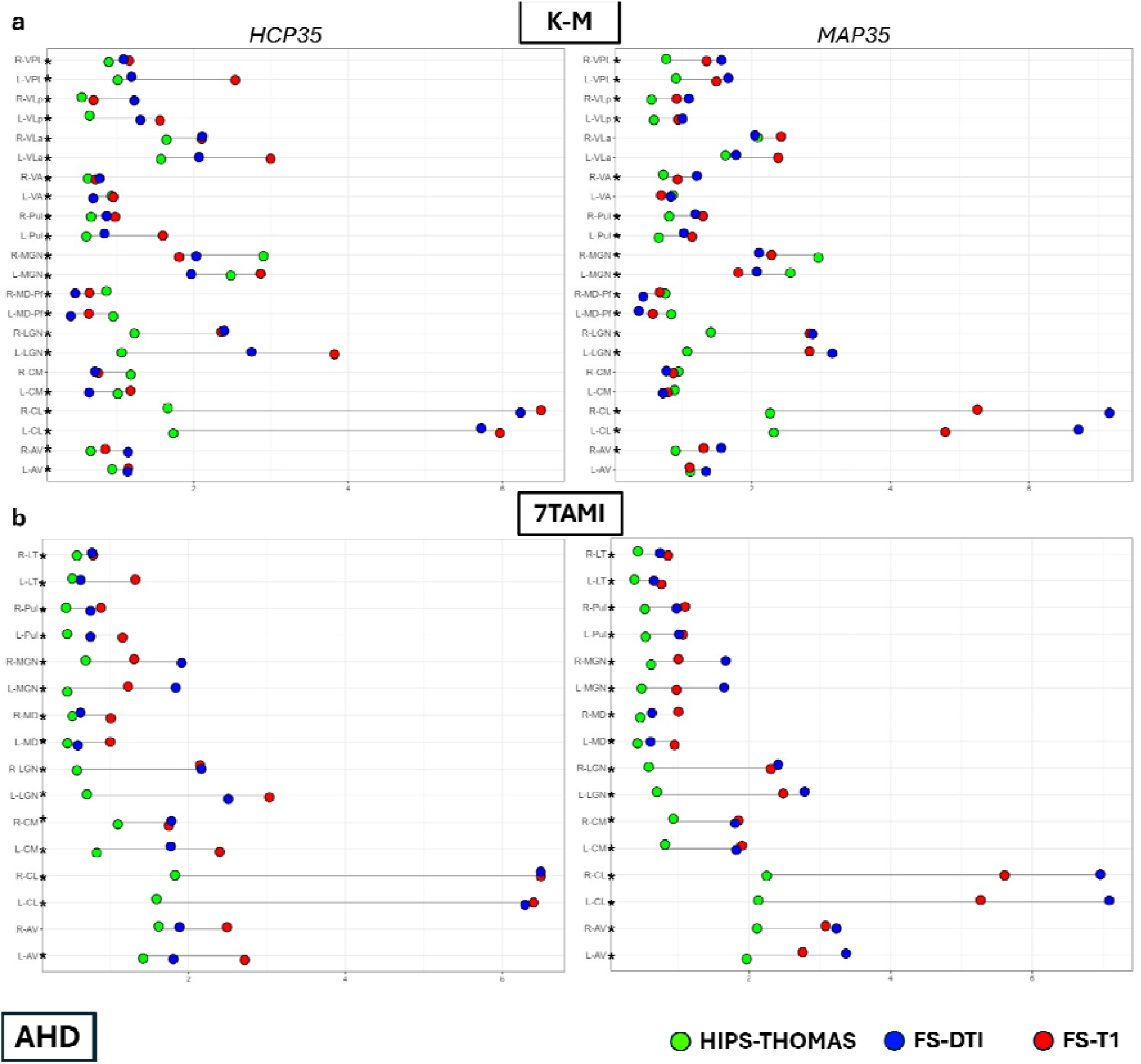
Dumbbell plots of AHD for nuclei segmented with FS-T1, FS-DTI, and HIPS-THOMAS relative to a) K-M and b) 7TAMI ground-truth reference segmentations (native space; see violin plots in Supplementary Figure 2); **key:** *: significantly lower AHD for one method relative to the other two; AV: anteroventral nucleus; LT: lateral nuclei (7TAMI); VA: ventral anterior nucleus; VLa: Ventrolateral anterior nucleus; VLp: Ventrolateral posterior nucleus; MD-Pf: mediodorsal-parafascicular nuclei; Pul: pulvinar nucleus; VPL: Ventral Posterolateral nucleus; CL: Centrolateral nucleus; CM: Centromedian nucleus; LGN: Lateral Geniculate Nucleus; MGN: Medial Geniculate Nucleus; L/R: Left, Right hemisphere; FS-T1: T_1_-based FreeSurfer segmentation (Iglesias et al., 2018); FS-DTI: FreeSurfer’s joint segmentation of thalamic nuclei from T1 scan and DTI (Tregidgo et al., 2023); HIPS-THOMAS: Thalamus Optimized Multi-atlas Segmentation using Histogram-based Polynomial Synthesis (Vidal et al., 2024); AHD: Average Hausdorff Distance; MAP35, HCP35: datasets with T1-weighted MRIs and diffusion MRI available; black diamond: mean; black horizontal line: median. K-M: Krauth-Morel atlas (Krauth et al., 2010); 7TAMI: the custom-made MNI version of the atlas of deep grey matter nuclei, part of the 7TAMIbrain dataset (Brun et al., 2022).

### MNI-space analysis

Group-level Dice coefficients and AHDs in MNI-space are tabulated in Figures 3 and 4, respectively. Overall, segmentations had higher Dice coefficients and lower AHDs along the diagonal (same K-M and 7TAMI segmentations) across segmentation methods. However, several exceptions were noted, indicating failure to distinguish between segmentations. These selectively occurred for FS-DTI and FS-T1. They involved L/R CL (relative to K-M’s AV, CM, MD-Pf, Pul, VA, VLa, VLp, and/or VPL, and 7TAMI’s AV, CM, and MD; notably, no voxels surpassed the probability threshold of 0.25 for the L/R CL segmentations for FS-T1 in the HCP35 dataset), L/R LGN (relative to K-M’s MGN, and/or Pul, and 7TAMI’s Pul), L/R VLa (relative to K-M’ s VLp and/or VA), and L VPL (relative to K-M’s L Pul).

**Figure 3:**
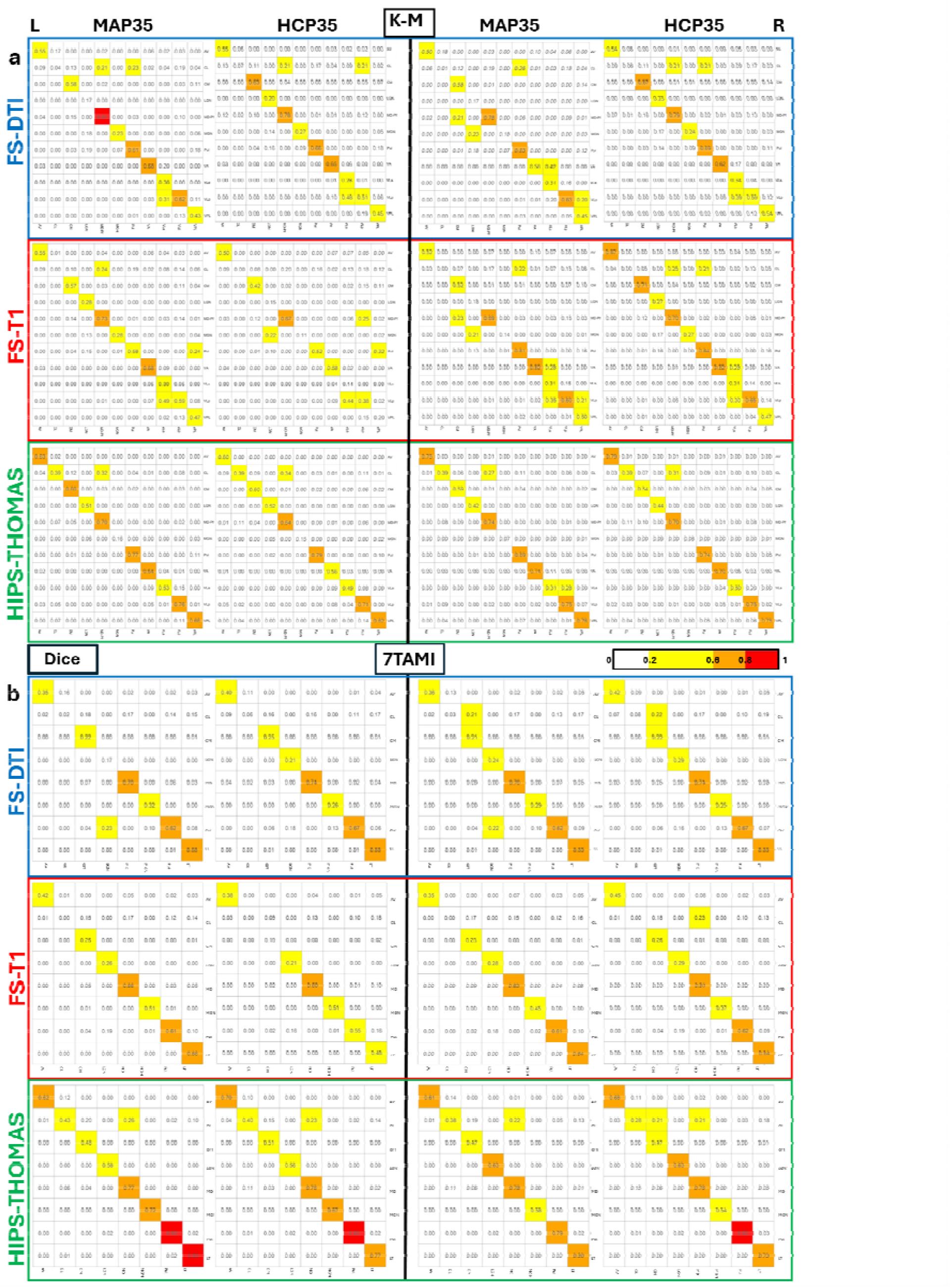
Group-level Dice coefficients for segmentations based on FS-T1, FS-DTI, and HIPS-THOMAS from MAP35, HCP35 (MNI space) relative to a) K-M and b) 7TAMI ground-truth reference segmentations: datasets with T1-weighted MRIs and diffusion MRI available**; key:** AV: anteroventral nucleus; LT: lateral nuclei (7TAMI); VA: ventral anterior nucleus; VLa: Ventrolateral anterior nucleus; VLp: Ventrolateral posterior nucleus; MD-Pf: mediodorsal-parafascicular nuclei; Pul: pulvinar nucleus; VPL: Ventral Posterolateral nucleus; CL: Centrolateral nucleus; CM: Centromedian nucleus; LGN: Lateral Geniculate Nucleus; MGN: Medial Geniculate Nucleus; L/R: Left, Right hemisphere; FS-T1: T_1_-based FreeSurfer segmentation (Iglesias et al., 2018); FS-DTI: FreeSurfer’s joint segmentation of thalamic nuclei from T1 scan and DTI (Tregidgo et al., 2023); HIPS-THOMAS: Thalamus Optimized Multi-atlas Segmentation using Histogram-based Polynomial Synthesis (Vidal et al., 2024); K-M: Krauth-Morel atlas (Krauth et al., 2010); 7TAMI: the custom-made MNI version of the atlas of deep grey matter nuclei, part of the 7TAMIbrain dataset (Brun et al., 2022).

**Figure 4:**
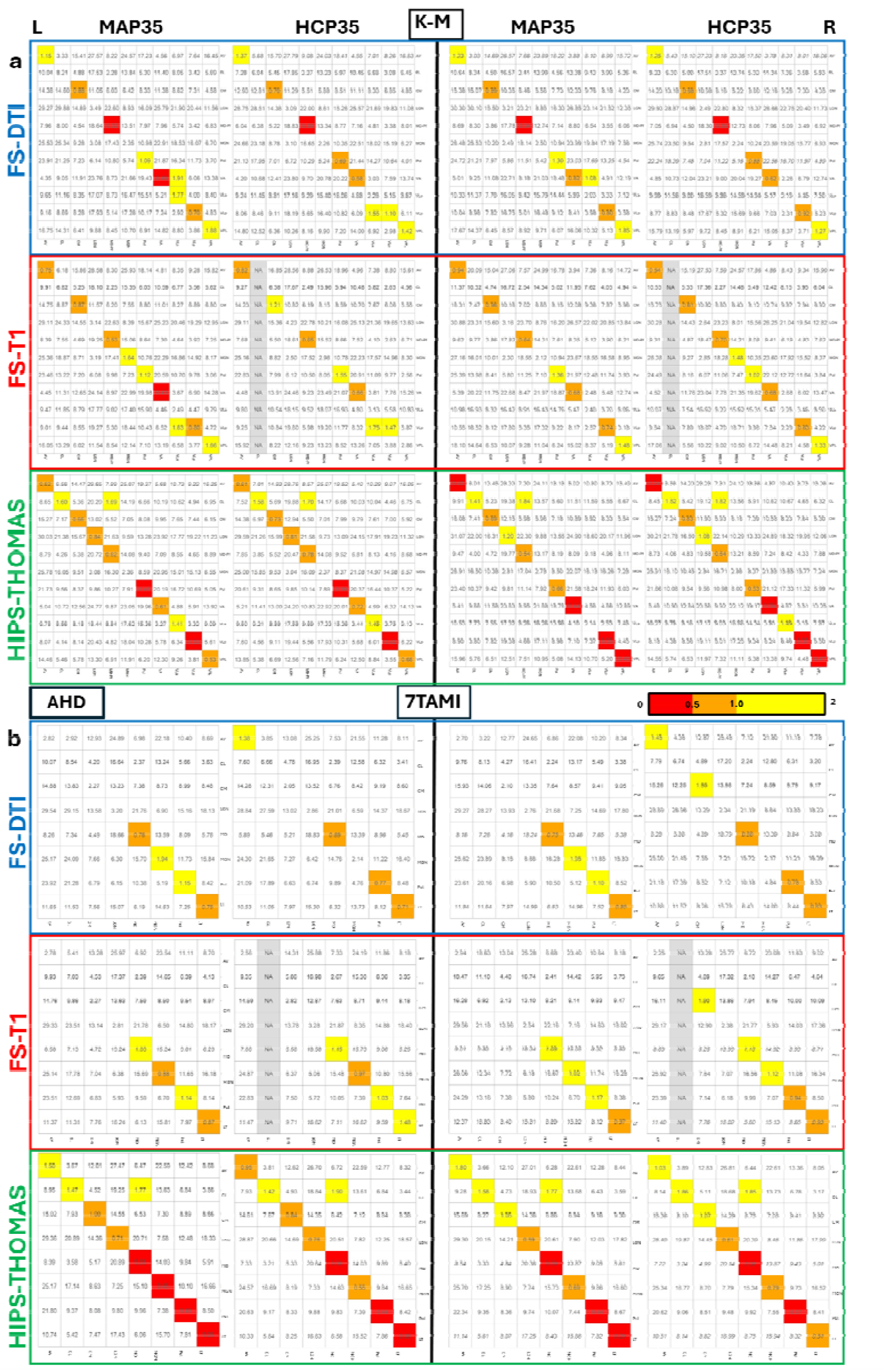
Group-level AHDs for segmentations based on FS-T1, FS-DTI, and HIPS-THOMAS from MAP35, HCP35 (MNI space) relative to a) K-M and b) 7TAMI ground-truth reference segmentations; **key:** AV: anteroventral nucleus; LT: lateral nuclei (7TAMI); VA: ventral anterior nucleus; VLa: Ventrolateral anterior nucleus; VLp: Ventrolateral posterior nucleus; MD-Pf: mediodorsal-parafascicular nuclei; Pul: pulvinar nucleus; VPL: Ventral Posterolateral nucleus; CL: Centrolateral nucleus; CM: Centromedian nucleus; LGN: Lateral Geniculate Nucleus; MGN: Medial Geniculate Nucleus; L/R: Left, Right hemisphere; FS-T1: T_1_-based FreeSurfer segmentation (Iglesias et al., 2018); FS-DTI: FreeSurfer’s joint segmentation of thalamic nuclei from T1 scan and DTI (Tregidgo et al., 2023); HIPS-THOMAS: Thalamus Optimized Multi-atlas Segmentation using Histogram-based Polynomial Synthesis (Vidal et al., 2024); AHD: Average Hausdorff Distance; K-M: Krauth-Morel atlas (Krauth et al., 2010); 7TAMI: the custom-made MNI version of the atlas of deep grey matter nuclei, part of the 7TAMIbrain dataset (Brun et al., 2022).

### Nucleus-wise “best” segmentation method

Overall, for both native-space and MNI-space analyses, for both the HCP35 and the MAP35 datasets, and for both ground-truth atlases consulted, HIPS-THOMAS consistently showed a greater number of segmentations with a Dice coefficient of ≥0.6 (at least “substantial” agreement/overlap) and an AHD≤1 (sufficiently small distance) than both FS-DTI and FS-T1. That was not consistently the case for FS-DTI relative to FS-T1 (Supplementary Table 8).

Using the K-M atlas as reference, HIPS-THOMAS emerged as the “best” of the three methods for 12/22 nuclei (R AV, L/R CL, L/R LGN, L/R Pul, L/R VLp, L/R VPL; Fig. 5a). FS-DTI was the “best” for only 2/22 nuclei (L/R MD-Pf). FS-T1 did not surpass the other two methods in any segmentation (0/22). FS-T1 and FS-DTI were joint-best for the R MGN. Ignoring HIPS-THOMAS, FS-DTI was consistently better (with respect to 8/8 criteria per segmentation) than FS-T1 only for 4/22 segmentations (L/R MD-Pf, L Pul, L VLa). HIPS-THOMAS was closest to being the best for L AV (5/8 criteria satisfied) and L VLa (7/8), whereas FS-DTI was closest to being the best only for L CM (5/8). Using 7TAMI as reference, FS-T1 and FS-DTI attained no “best” status for any of the 16 segmentations. HIPS-THOMAS was the best (8/8 criteria satisfied) for 11/16 nuclei, and closest to being the best for the remaining 5/16, satisfying 7/8 criteria for all 5 nuclei.

**Figure 5.**
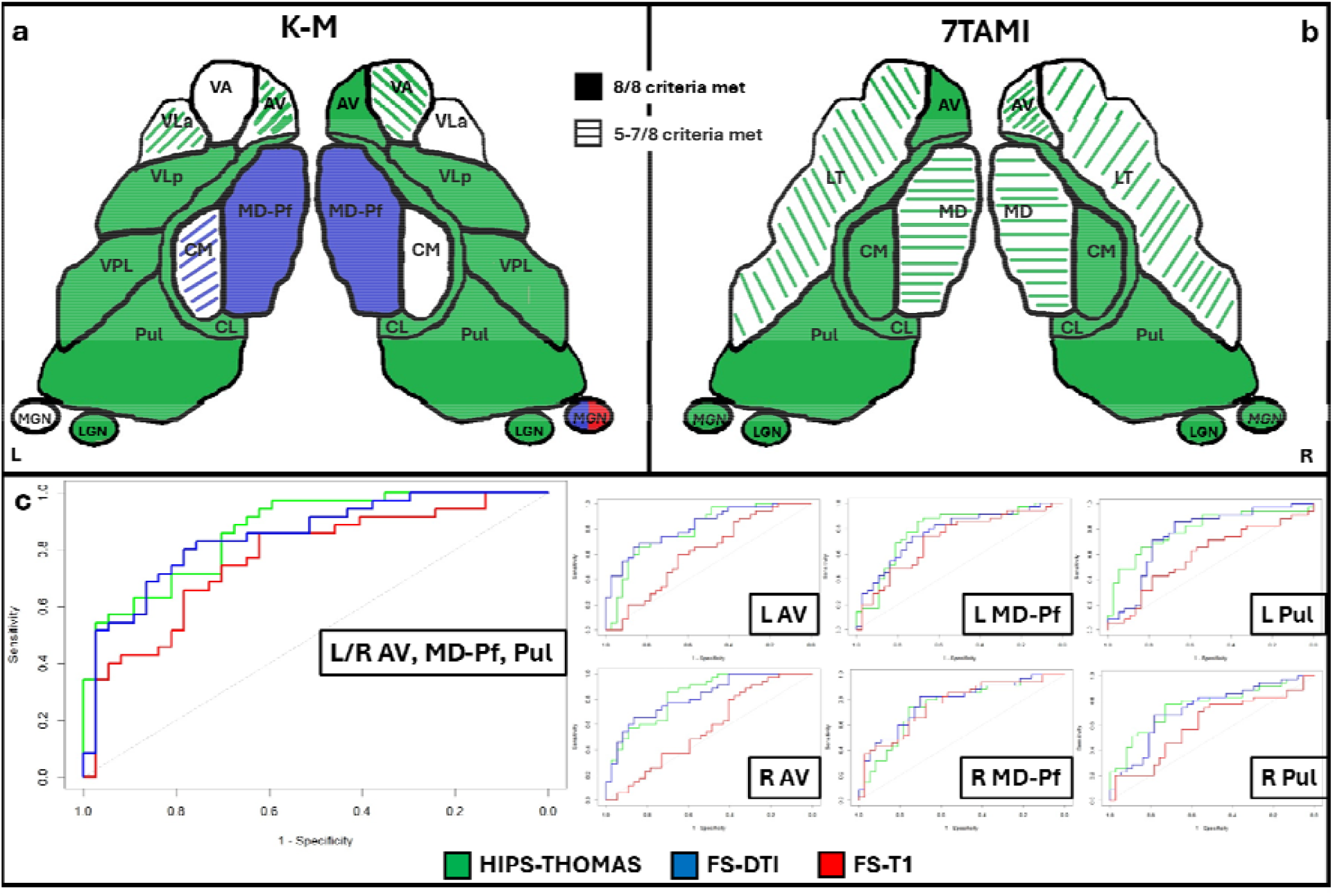
illustration of the best segmentation method for the 22 nuclei examined relative to a) K-M and b) 7TAMI; solid fill: best segmentation method; pattern fill (dashed lines): close-to-best method; no fill: no best or close-to-best method identified; c: ROC curves illustrating the three separate binomial logistic regression analyses (one per segmentation method), using all 6 segmentations of interest; c: ROC curves illustrating the 18 binomial logistic regression analyses conducted separately for each nucleus and segmentation method; **key:** AV: anteroventral nucleus; VA: ventral anterior nucleus; VLa: Ventrolateral anterior nucleus; LT: lateral nuclei (7TAMI); VLp: Ventrolateral posterior nucleus; MD-Pf: mediodorsal-parafascicular nuclei; Pul: pulvinar nucleus; VPL: Ventral Posterolateral nucleus; CL: Centrolateral nucleus; CM: Centromedian nucleus; LGN: Lateral Geniculate Nucleus; MGN: Medial Geniculate Nucleus; L/R: Left, Right hemisphere; FS-T1: T_1_-based FreeSurfer segmentation (Iglesias et al., 2018); FS-DTI: FreeSurfer’s joint segmentation of thalamic nuclei from T1w-MRI and DTI (Tregidgo et al., 2023); HIPS-THOMAS: Thalamus Optimized Multi-atlas Segmentation using Histogram-based Polynomial Synthesis (Vidal et al., 2024); K-M: Krauth-Morel atlas (Krauth et al., 2010); 7TAMI: the custom-made MNI version of the atlas of deep grey matter nuclei, part of the 7TAMIbrain dataset (Brun et al., 2022).

### ROC analysis

In three (one per segmentation method) logistic binomial regression analyses including all 6 volumes of interest as predictors (L/R AV, MD-Pf, Pul), HIPS-THOMAS showed numerically the largest AUC (0.868) relative to FS-DTI (0.847) and FS-T1 (0.768; Fig. 5b). DeLong comparisons showed that FS-T1’s AUC was only marginally smaller than those of HIPS-THOMAS (z = 2.10, p-corr(3) = 0.107) and FS-DTI (z = 1.73, p-corr(3) = 0.166). AUC differences between HIPS-THOMAS and FS-DTI did not approach significance (z = 0.51, p-corr(3) = 0.608).

A series of 18 analyses (6 volumes * 3 methods) using each volume as a single predictor, showed that L and R MD-Pf volumes were significant (p-corr < 0.05) predictors of group membership (control vs patient) for all three methods. However, L/R AV and Pul volumes were significant predictors only for HIPS-THOMAS and FS-DTI (Table 1; Fig. 5c). DeLong comparisons (n=18) between different methods on each of those 6 volumes, showed no difference amongst the three methods with respect to AUCs for MD-Pf, or between HIPS-THOMAS and FS-DTI on any of the 6 volumes. However, FS-T1’s AUCs were significantly or marginally smaller than those of HIPS-THOMAS and FS-DTI for L/R AV and Pul (Table 2).

## Discussion

Recent developments in automated segmentation of the human thalamus have substantially facilitated research on the integrity, activity, and connectivity of individual thalamic nuclei in health and disease. Although there are currently several methods available, studies comparing their accuracy and reliability are scant. Relying on suboptimal segmentation methods and cherry-picking amongst alternatives may compromise the efficacy of neuromodulatory interventions targeting the thalamus, as well as the replication of comparative neuroanatomical studies and clinical cognitive neuroimaging research.

Here, we used two datasets of healthy adults with T1w-MRI and diffusion MRI data to compare amongst three state-of-the art structural MRI-based thalamic segmentation methods: the more widely used one (FS-T1; Iglesias et al., 2018), along with two more recently developed ones: FS-DTI (Tregidgo et al., 2023), and HIPS-THOMAS (Vidal et al., 2024). Unlike the other two methods, FS-DTI leverages information from both T1w-MRI and diffusion MRI. Whereas HIPS-THOMAS has been shown to surpass FS-T1 (Williams et al., 2024), it has not been compared with FS-DTI, and we are also unaware of the extent to which FS-DTI enhances thalamic segmentation relative to FS-T1. Moreover, some of the work that has highlighted limitations in FS-T1 (Williams et al., 2022, 2024) has relied on the K-M atlas (Jakab et al., 2012; Krauth et al., 2010) as a silver-standard ground-truth – the use of which as such is not unproblematic (Brun et al., 2022). It is thus important to examine whether comparisons between segmentation methods can be replicated with alternative reference atlases, other than the K-M atlas.

Surprisingly, despite relying exclusively on structural T1w-MRI inputs, HIPS-THOMAS consistently outperformed, not just FS-T1 (dovetailing with earlier observations in Williams et al., 2024), but also FS-DTI, and did so irrespective of the ground-truth atlas used - 7TAMI or K-M. Using the K-M, HIPS-THOMAS outperformed the other two on 12/22 segmentations (R AV, R VA, L/R CL, L/R LGN, L/R Pul, L/R VLp, L/R VPL) and outperformed them in a less consistent fashion in another 2 (L AV and L VLa). FS-DTI consistently outperformed FS-T1 in only 4/22 segmentations (L/R MD-Pf, L Pul, L VLa) when using K-M, and in 3/16 (L/R MD-Pf, R Pul) when using 7TAMI. Using the 7TAMI, HIPS-THOMAS outperformed the other two on all 16 segmentations – consistently (8/8 criteria met) on 11/16, and less consistently (7/8 criteria met) on the remaining 5/16 (L/R MD-Pf, L/R V, R AV).

When using the K-M atlas, the only segmentation in which FS-DTI outperformed HIPS-THOMAS and FS-T1 was the MD-Pf, bilaterally, and less consistently for L CM. Even so, across datasets and analyses (mean values for native space, group MNI values), the HIPS-THOMAS MD-Pf segmentations showed Dice coefficients that had a minimum of 0.58, and AHDs with a maximum of 0.96. Moreover, in the AUC analyses, the MD-Pf volumes from HIPS-THOMAS remained significant predictors of group membership and did not differ in this regard from FS-T1 or FS-DTI. However, HIPS-THOMAS did not show these limitations when using 7TAMI as the ground truth. Given the inconsistencies between the two ground truths on MD-Pf, CM, and MGN, it would be important to approach the findings reported in Williams et al. (2024) on these segmentations with caution. In that paper, FS-T1 was the best segmentation method for the L MD-Pf, the R CM (and joint best with HIPS-THOMAS on the L CM), and for L MGN. Whether this should be attributed to the limitations of the K-M atlas is outside the scope of our investigation. Moreover, given that HIPS-THOMAS segmentations are substantially fewer than those of FS-T1/FS-DTI, we have confined our analyses to segmentations that are common across all three of the methods. Our comments on the improvement that FS-DTI makes relative to FS-T1 thus pertain to only those 22 nuclei in question.

Even so, FS-T1’s failure to discriminate between healthy controls and patients on the grounds of AV and Pul volumes is strongly consistent with Williams et al. (2024), wherein FS-T1 performed quite poorly in discriminating healthy controls from those with mild cognitive impairment and Alzheimer’s disease for AV and Pul (*ibid*, Fig. 7; note that FS-DTI was not part of these comparisons). Given that automated thalamic segmentation may be affected by condition-specific abnormalities in neurological patients (e.g., ventricular enlargement, white matter damage in tracts connecting the thalamus; Segobin et al., 2024), comparisons of these methods in different conditions are particularly important. Along with those of Williams et al. (2024), our AUC analyses thus highlight the danger of false negatives in studies that have relied on FS-T1.

Arguably, low-quality segmentations may also result in false positives. Several studies using FS-T1 have disclosed volume reduction in segmentations that we and Williams et al. (2024) have shown to have poor segmentation accuracy. Examples include the reduction observed (among others) in AV, LGN, VA, VLa, VLp, and VPL in major depressive disorder (Zhang et al., 2024), in Pul for pediatric bipolar disorder (Gao et al., 2025), in Pul and LGN for prenatal alcohol exposure (Nakhid et al., 2023), in Pul for remitting-relapsing multiple sclerosis (Trufanov et al., 2021), in Pul and LGN for schizophrenia spectrum disorders and bipolar disorders (Mørch-Johnsen et al., 2023). Likewise, caution would be advisable in interpreting the findings of tractography studies using FS-T1 to segment the aforementioned nuclei - e.g., LGN and VLp in Liu et al. (2022).

To potentially minimise measurement error along with the number of statistical tests, several studies (e.g., Bergsland et al., 2021; McKenna et al., 2022; Pardilla-Delgado et al., 2021; Shah et al., 2024; Thalhammer et al., 2024; Tung et al., 2022) that employ FS-T1, group thalamic nuclei into subregions, based on their spatial arrangement (e.g., the AV is assigned to the anterior region, the LD and LP assigned to a lateral region, the LGN, MGN, LSg, PuA, PuM, PuL, and PuI may be assigned to a posterior region), or their functional anatomy (e.g., the AV may be its own group, whereas the VA, VAmc, VLa, and VLp may be assigned to a ‘Motor’ group). Apart from impeding the localisation of the effects of interest, this approach ignores the substantial variability across these composite ROIs with respect to their volume, the number of nuclei each of those comprises, and, as shown here, their segmentation quality.

Finally, the fact that HIPS-THOMAS was as good as FS-DTI in distinguishing between healthy controls and patients on the basis of the volumes of nuclei of interest is noteworthy. Substantial resources are involved in collecting the data necessary (diffusion MRI) and completing the additional preprocessing stages to derive the FS-DTI segmentations. The use of FS-DTI would be further impeded by the fact that several open MRI databases may contain T1w-MRI, but not high-quality diffusion MRI data. On the other hand, HIPS-THOMAS holds the additional advantage of catering for the segmentation of both thalamic nuclei and other subcortical structures with much shorter computational time and within a single unified package - see discussion in Saranathan et al. (2025).

Given the above, we strongly urge the community to exercise caution whilst using FS-T1, revert to HIPS-THOMAS when possible, and, depending on the availability of diffusion MRI data, corroborate findings with FS-DTI. Replicating FS-T1-based analyses in already published studies with HIPS-THOMAS and FS-DTI would also be instructive, for the purposes of minimising false positives/negatives.

## Data Availability

Demographic and volumetric data are publicly available on https://osf.io/8bp64/. Bash, Matlab, and R scripts are also available there. HCP35 (MGH HCP Adult Diffusion Dataset ID: MGH_DIFF): https://db.humanconnectome.org/data/projects/MGH_DIFF. Data collection and sharing for this project was provided by the Human Connectome Project (HCP; Principal Investigators: Bruce Rosen, M.D., Ph.D., Martinos Center at Massachusetts General Hospital; Arthur W. Toga, Ph.D., University of Southern California; Van J. Weeden, MD, Martinos Center at Massachusetts General Hospital). HCP funding was provided by the National Institute of Dental and Craniofacial Research (NIDCR), the National Institute of Mental Health (NIMH), and the National Institute of Neurological Disorders and Stroke (NINDS). HCP data are disseminated by the Laboratory of Neuro Imaging at the University of Southern California. HCP is the result of efforts of co-investigators from the University of Southern California, Martinos Center for Biomedical Imaging at Massachusetts General Hospital (MGH), Washington University, and the University of Minnesota. FS-T1: FreeSurfer segmentation was Conducted following the instructions provided in https://freesurfer.net/fswiki/ThalamicNuclei). HIPS-THOMAS: Details for running HIPS-THOMAS using Docker are available on Github (HIPS-THOMAS https://github.com/thalamicseg/hipsthomasdocker); the Docker images are available on
Docker Hub (https://hub.docker.com/u/anagrammarian). FS-DTI: details on its use can be found in https://surfer.nmr.mgh.harvard.edu/fswiki/ThalamicNucleiDTI. We used the updated model (`thalseg_1.1.h5`) made available https://ftp.nmr.mgh.harvard.edu/pub/dist/freesurfer/thalseg/ThalsegPatch_1.1.tgz) (validated against additional datasets) which we called with the -model option. Improvements to the segmentation script were downloaded from https://github.com/freesurfer/freesurfer/blob/dev/mri_segment_thalamic_nuclei_dti_cnn/mri_segment_thalamic_nuclei_dti_cnn and were used to replace the release version, in order to prevent occurrences of false positive thalamic segmentations; K-M atlas: available on https://zenodo.org/records/13918589; 7TAMIbrainDGN: available on https://github.com/arnaudletroter/7TAMIBrain

https://db.humanconnectome.org/data/projects/MGH_DIFF

https://freesurfer.net/fswiki/ThalamicNuclei

https://github.com/thalamicseg/hipsthomasdocker

https://hub.docker.com/u/anagrammarian

https://surfer.nmr.mgh.harvard.edu/fswiki/ThalamicNucleiDTI

https://ftp.nmr.mgh.harvard.edu/pub/dist/freesurfer/thalseg/ThalsegPatch_1.1.tgz

https://github.com/freesurfer/freesurfer/blob/dev/mri_segment_thalamic_nuclei_dti_cnn/mri_segment_thalamic_nuclei_dti_cnn

https://zenodo.org/records/13918589

https://github.com/arnaudletroter/7TAMIBrain

## Acknowledgements

- We are very grateful to the participants who took part in the Memory and Amnesia Project.
- We also thank Drs Clare Loane, Adriana Roca-Fernandez, Carmen Lage-Martinez, and Sarosh R Irani, for participant recruitment and data acquisition (Argyropoulos et al., 2019).
- CRB was supported by a Medical Research Council Clinician Scientist Fellowship (MR/K010395/1). The funders had no role in study design, data collection and interpretation, or the decision to submit the work for publication.
- MS acknowledges funding from the National Institute of Biomedical Imaging and Bioengineering (R01 award EB032674).

## Ethics

Memory and Amnesia Project: All participants provided written informed consent according to the Declaration of Helsinki. Ethical approval was received from South Central Oxford Research Ethics Committee (REC no: 08/H0606/133).

## Contributions / CRediT author statement

- G.P.D.A.: Conceptualization, Data curation, Formal analysis, Investigation, Visualization, Methodology, Writing – original draft, Writing – review & editing, Project administration
- C.R.B.: Funding acquisition, Supervision, Writing – original draft, Writing – review & editing, Project administration
- M.S.: Conceptualisation, Supervision, Methodology, Writing – original draft, Writing review & editing

## Data and code availability

**Demographic and volumetric data** are publicly available on https://osf.io/8bp64/. **Bash, Matlab, and R** scripts are also available there. **HCP35** (MGH HCP Adult Diffusion Dataset ID: “MGH_DIFF”): https://db.humanconnectome.org/data/projects/MGH_DIFF. Data collection and sharing for this project was provided by the Human Connectome Project (HCP; Principal Investigators: Bruce Rosen, M.D., Ph.D., Martinos Center at Massachusetts General Hospital; Arthur W. Toga, Ph.D., University of Southern California; Van J. Weeden, MD, Martinos Center at Massachusetts General Hospital). HCP funding was provided by the National Institute of Dental and Craniofacial Research (NIDCR), the National Institute of Mental Health (NIMH), and the National Institute of Neurological Disorders and Stroke (NINDS). HCP data are disseminated by the Laboratory of Neuro Imaging at the University of Southern California. HCP is the result of efforts of co-investigators from the University of Southern California, Martinos Center for Biomedical Imaging at Massachusetts General Hospital (MGH), Washington University, and the University of Minnesota. **FS-T1:** FreeSurfer segmentation was Conducted following the instructions provided in https://freesurfer.net/fswiki/ThalamicNuclei). **HIPS-THOMAS:** Details for running HIPS-THOMAS using Docker are available on Github (HIPS-THOMAS https://github.com/thalamicseg/hipsthomasdocker); the Docker images are available on Docker Hub (https://hub.docker.com/u/anagrammarian). **FS-DTI:** details on its use can be found in https://surfer.nmr.mgh.harvard.edu/fswiki/ThalamicNucleiDTI. We used the updated model (`thalseg_1.1.h5`) made available https://ftp.nmr.mgh.harvard.edu/pub/dist/freesurfer/thalseg/ThalsegPatch_1.1.tgz) (validated against additional datasets) which we called with the “—model” option. Improvements to the segmentation script were downloaded from https://github.com/freesurfer/freesurfer/blob/dev/mri_segment_thalamic_nuclei_dti_cnn/mri_segment_thalamic_nuclei_dti_cnn and were used to replace the release version, in order to prevent occurrences of false positive thalamic segmentations; **K-M atlas**: available on https://zenodo.org/records/13918589; **7TAMIbrainDGN:** available on https://github.com/arnaudletroter/7TAMIBrain

## References

Andersson, J., Smith, S., & Jenkinson, M. (2008, 19/06). FNIRT-FMRIB’s non-linear image registration tool. 14th Annual Meeting of the Organization for Human Brain Mapping, Melbourne, Victoria, Australia.

Argyropoulos, G. P. D., Aggleton, J. P., & Butler, C. R. (2025). Diencephalic and white matter knock-on effects in hippocampal amnesia – why they matter (p. 2025.05.21.655297). bioRxiv. 10.1101/2025.05.21.655297

Argyropoulos, G. P. D., Loane, C., Roca-Fernandez, A., Lage-Martinez, C., Gurau, O., Irani, S. R., & Butler, C. R. (2019). Network-wide abnormalities explain memory variability in hippocampal amnesia. eLife, 8. 10.7554/eLife.46156

Battistella, G., Najdenovska, E., Maeder, P., Ghazaleh, N., Daducci, A., Thiran, J.-P., Jacquemont, S., Tuleasca, C., Levivier, M., Bach Cuadra, M., & Fornari, E. (2017). Robust thalamic nuclei segmentation method based on local diffusion magnetic resonance properties. Brain Structure and Function, 222(5), 2203–2216. 10.1007/s00429-016-1336-4

Bergsland, N., Benedict, R. H. B., Dwyer, M. G., Fuchs, T. A., Jakimovski, D., Schweser, F., Tavazzi, E., Weinstock-Guttman, B., & Zivadinov, R. (2021). Thalamic Nuclei Volumes and Their Relationships to Neuroperformance in Multiple Sclerosis: A Cross-Sectional Structural MRI Study. Journal of Magnetic Resonance Imaging, 53(3), 731–739. 10.1002/jmri.27389

Brun, G., Testud, B., Girard, O. M., Lehmann, P., de Rochefort, L., Besson, P., Massire, A., Ridley, B., Girard, N., Guye, M., Ranjeva, J.-P., & Le Troter, A. (2022). Automatic segmentation of deep grey nuclei using a high-resolution 7T magnetic resonance imaging atlas—Quantification of T1 values in healthy volunteers. European Journal of Neuroscience, 55(2), 438–460. 10.1111/ejn.15575

Gao, W., Zhang, K., Jiao, Q., Su, L., Cui, D., Lu, S., & Yang, R. (2025). Machine learning for classification of pediatric bipolar disorder with and without psychotic symptoms based on thalamic subregional structural volume. BMC Psychiatry, 25(1), 574. 10.1186/s12888-025-07018-5

Gentile, C. P., Spitschan, M., Taskin, H. O., Bock, A. S., & Aguirre, G. K. (2024). Temporal Sensitivity for Achromatic and Chromatic Flicker across the Visual Cortex. Journal of Neuroscience, 44(21). 10.1523/JNEUROSCI.1395-23.2024

Holm, S. (1979). A simple sequentially rejective multiple test procedure. Scandinavian Journal of Statistics, 6(6), 65–70. 10.2307/4615733

Iglehart, C., Monti, M., Cain, J., Tourdias, T., & Saranathan, M. (2020). A systematic comparison of structural-, structural connectivity-, and functional connectivity-based thalamus parcellation techniques. Brain Structure and Function, 225(5), 1631–1642. 10.1007/s00429-020-02085-8

Iglesias, J. E., Insausti, R., Lerma-Usabiaga, G., Bocchetta, M., Van Leemput, K., Greve, D. N., van der Kouwe, A., Fischl, B., Caballero-Gaudes, C., & Paz-Alonso, P. M. (2018). A probabilistic atlas of the human thalamic nuclei combining ex vivo MRI and histology. NeuroImage, 183, 314–326. 10.1016/j.neuroimage.2018.08.012

Jakab, A., Blanc, R., Berényi, E. L., & Székely, G. (2012). Generation of individualized thalamus target maps by using statistical shape models and thalamocortical tractography. American Journal of Neuroradiology, 33(11). 10.3174/ajnr.A3140

Jenkinson, M., Bannister, P., Brady, M., & Smith, S. (2002). Improved Optimization for the Robust and Accurate Linear Registration and Motion Correction of Brain Images. NeuroImage, 17(2), 825–841. 10.1006/nimg.2002.1132

Krauth, A., Blanc, R., Poveda, A., Jeanmonod, D., Morel, A., & Székely, G. (2010). A mean three-dimensional atlas of the human thalamus: Generation from multiple histological data. NeuroImage, 49(3), 2053–2062. 10.1016/j.neuroimage.2009.10.042

Lee, H.-J., Seo, S. A., Lee, B. I., Kim, S. E., & Park, K. M. (2020). Thalamic nuclei volumes and network in juvenile myoclonic epilepsy. Acta Neurologica Scandinavica, 141(4), 271– 278. 10.1111/ane.13198

Liu, M., Lerma-Usabiaga, G., Clascá, F., & Paz-Alonso, P. M. (2022). Reproducible protocol to obtain and measure first-order relay human thalamic white-matter tracts. NeuroImage, 262, 119558. 10.1016/j.neuroimage.2022.119558

Maffei, C., Lee, C., Planich, M., Ramprasad, M., Ravi, N., Trainor, D., Urban, Z., Kim, M., Jones, R. J., Henin, A., Hofmann, S. G., Pizzagalli, D. A., Auerbach, R. P., Gabrieli, J. D. E., Whitfield-Gabrieli, S., Greve, D. N., Haber, S. N., & Yendiki, A. (2021). Using diffusion MRI data acquired with ultra-high gradient strength to improve tractography in routine-quality data. NeuroImage, 245, 118706. 10.1016/j.neuroimage.2021.118706

McKenna, M. C., Li Hi Shing, S., Murad, A., Lope, J., Hardiman, O., Hutchinson, S., & Bede, P. (2022). Focal thalamus pathology in frontotemporal dementia: Phenotype-associated thalamic profiles. Journal of the Neurological Sciences, 436, 120221. 10.1016/j.jns.2022.120221

Mørch-Johnsen, L., Jørgensen, K. N., Barth, C., Nerland, S., Bringslid, I. K., Wortinger, L. A., Andreou, D., Melle, I., Andreassen, O. A., & Agartz, I. (2023). Thalamic nuclei volumes in schizophrenia and bipolar spectrum disorders – Associations with diagnosis and clinical characteristics. Schizophrenia Research, 256, 26–35. 10.1016/j.schres.2023.04.008

Morel, A. (2007). Stereotactic Atlas of the Human Thalamus and Basal Ganglia. CRC Press.

Morel, A., Magnin, M., & Jeanmonod, D. (1997). Multiarchitectonic and stereotactic atlas of the human thalamus. Journal of Comparative Neurology, 387(4), 588–630. 10.1002/(SICI)1096-9861(19971103)387:4%253C588::AID-CNE8%253E3.0.CO;2-Z

Najdenovska, E., Alemán-Gómez, Y., Battistella, G., Descoteaux, M., Hagmann, P., Jacquemont, S., Maeder, P., Thiran, J.-P., Fornari, E., & Bach Cuadra, M. (2018). In-vivo probabilistic atlas of human thalamic nuclei based on diffusion-weighted magnetic resonance imaging. Scientific Data, 5(1), 180270. 10.1038/sdata.2018.270

Nakhid, D., Patel, D., McMorris, C. A., Gibbard, W. B., Tortorelli, C., Pei, J., & Lebel, C. (2023). Limbic brain subregions associated with mental health symptoms in youth with and without prenatal alcohol exposure. Alcohol, Clinical and Experimental Research, 47(11), 2033–2044. 10.1111/acer.15181

Pajula, J., Kauppi, J.-P., & Tohka, J. (2012). Inter-Subject Correlation in fMRI: Method Validation against Stimulus-Model Based Analysis. PLOS ONE, 7(8), e41196. 10.1371/journal.pone.0041196

Pardilla-Delgado, E., Torrico-Teave, H., Sanchez, J. S., Ramirez-Gomez, L. A., Baena, A., Bocanegra, Y., Vila-Castelar, C., Fox-Fuller, J. T., Guzmán-Vélez, E., Martínez, J., Alvarez, S., Ochoa-Escudero, M., Lopera, F., & Quiroz, Y. T. (2021). Associations between subregional thalamic volume and brain pathology in autosomal dominant Alzheimer’s disease. Brain Communications, 3(2), fcab101. 10.1093/braincomms/fcab101

Saranathan, M., Cogliandro, G., Hicks, T., Patterson, D., Vachha, B., & Cacciola, A. (2025). Comprehensive segmentation of deep grey nuclei from structural MRI data (No. 2503.21955). arXiv. 10.48550/arXiv.2503.21955

Segobin, S., Haast, R. A. M., Kumar, V. J., Lella, A., Alkemade, A., Bach Cuadra, M., Barbeau, E. J., Felician, O., Pergola, G., Pitel, A.-L., Saranathan, M., Tourdias, T., & Hornberger, M. (2024). A roadmap towards standardized neuroimaging approaches for human thalamic nuclei. Nature Reviews Neuroscience, 25(12), 792–808. 10.1038/s41583-024-00867-1

Shah, S. N., Dounavi, M.-E., Malhotra, P. A., Lawlor, B., Naci, L., Koychev, I., Ritchie, C. W., Ritchie, K., & O’Brien, J. T. (2024). Dementia risk and thalamic nuclei volumetry in healthy midlife adults: The PREVENT Dementia study. Brain Communications, 6(2), fcae046. 10.1093/braincomms/fcae046

Shin, K. J., Lee, H.-J., & Park, K. M. (2019). Alterations of individual thalamic nuclei volumes in patients with migraine. The Journal of Headache and Pain, 20(1), 112. 10.1186/s10194-019-1063-3

Su, J. H., Thomas, F. T., Kasoff, W. S., Tourdias, T., Choi, E. Y., Rutt, B. K., & Saranathan, M. (2019). Thalamus Optimized Multi Atlas Segmentation (THOMAS): Fast, fully automated segmentation of thalamic nuclei from structural MRI. NeuroImage, 194, 272–282. 10.1016/j.neuroimage.2019.03.021

Taha, A. A., & Hanbury, A. (2015). Metrics for evaluating 3D medical image segmentation: Analysis, selection, and tool. BMC Medical Imaging, 15(1), 29. 10.1186/s12880-015-0068-x

Thalhammer, M., Schulz, J., Scheulen, F., Oubaggi, M. E. M., Kirschner, M., Kaiser, S., Schmidt, A., Borgwardt, S., Avram, M., Brandl, F., & Sorg, C. (2024). Distinct Volume Alterations of Thalamic Nuclei Across the Schizophrenia Spectrum. Schizophrenia Bulletin, 50(5), 1208–1222. 10.1093/schbul/sbae037

Tregidgo, H. F. J., Soskic, S., Olchanyi, M. D., Althonayan, J., Billot, B., Maffei, C., Golland, P., Yendiki, A., Alexander, D. C., Bocchetta, M., Rohrer, J. D., & Iglesias, J. E. (2023). Domain-agnostic segmentation of thalamic nuclei from joint structural and diffusion MRI (Vol. 14227, pp. 247–257). 10.1007/978-3-031-43993-3_24

Trufanov, A., Bisaga, G., Skulyabin, D., Temniy, A., Poplyak, M., Chakchir, O., Efimtsev, A., Dmitriy, T., Odinak, M., & Litvinenko, I. (2021). Thalamic nuclei degeneration in multiple sclerosis. Journal of Clinical Neuroscience, 89, 375–380. 10.1016/j.jocn.2021.05.043

Tung, H., Pan, S.-Y., Lan, T.-H., Lin, Y.-Y., & Peng, S.-J. (2022). Characterization of Hippocampal-Thalamic-Cortical Morphometric Reorganization in Temporal Lobe Epilepsy. Frontiers in Neurology, 12, 810186. 10.3389/fneur.2021.810186

Umapathy, L., Keerthivasan, M. B., Zahr, N. M., Bilgin, A., & Saranathan, M. (2022). Convolutional Neural Network Based Frameworks for Fast Automatic Segmentation of Thalamic Nuclei from Native and Synthesized Contrast Structural MRI. Neuroinformatics, 20(3), 651–664. 10.1007/s12021-021-09544-5

van Oort, E. S. B., Mennes, M., Navarro Schröder, T., Kumar, V. J., Zaragoza Jimenez, N.I., Grodd, W., Doeller, C. F., & Beckmann, C. F. (2018). Functional parcellation using time courses of instantaneous connectivity. Segmenting the Brain, 170, 31–40. 10.1016/j.neuroimage.2017.07.027

Velioglu, H. A., Ayyildiz, B., Ayyildiz, S., Sutcubasi, B., Hanoglu, L., Bayraktaroglu, Z., & Yulug, B. (2023). A structural and resting-state functional connectivity investigation of the pulvinar in elderly individuals and Alzheimer’s disease patients. Alzheimer’s & Dementia, 19(7), 2774–2789. 10.1002/alz.12850

Vidal, J. P., Danet, L., Péran, P., Pariente, J., Bach Cuadra, M., Zahr, N. M., Barbeau, E. J., & Saranathan, M. (2024). Robust thalamic nuclei segmentation from T1-weighted MRI using polynomial intensity transformation. Brain Structure & Function. 10.1007/s00429-024-02777-5

Weeland, C. J., Vriend, C., van der Werf, Y., Huyser, C., Hillegers, M., Tiemeier, H., White, T., & van den Heuvel, O. A. (2022). Thalamic Subregions and Obsessive-Compulsive Symptoms in 2,500 Children From the General Population. Journal of the American Academy of Child & Adolescent Psychiatry, 61(2), 321–330. 10.1016/j.jaac.2021.05.024

Williams, B., Nguyen, D., Vidal, J. P., & Saranathan, M. (2024). Thalamic nuclei segmentation from T1-weighted MRI: Unifying and benchmarking state-of-the-art methods. Imaging Neuroscience, 2, 1–16. 10.1162/imag_a_00166

Williams, B., Roesch, E., & Christakou, A. (2022). Systematic validation of an automated thalamic parcellation technique using anatomical data at 3T. NeuroImage, 258, 119340. 10.1016/j.neuroimage.2022.119340

Xiong, G., Dong, D., Cheng, C., Jiang, Y., Sun, X., He, J., Li, C., Gao, Y., Zhong, X., Zhao, H., Wang, X., & Yao, S. (2021). Potential structural trait markers of depression in the form of alterations in the structures of subcortical nuclei and structural covariance network properties. NeuroImage: Clinical, 32, 102871. 10.1016/j.nicl.2021.102871

Yendiki, A., Panneck, P., Srinivasan, P., Stevens, A., Zöllei, L., Augustinack, J., Wang, R., Salat, D., Ehrlich, S., Behrens, T., Jbabdi, S., Gollub, R., & Fischl, B. (2011). Automated Probabilistic Reconstruction of White-Matter Pathways in Health and Disease Using an Atlas of the Underlying Anatomy. Frontiers in Neuroinformatics, 5. 10.3389/fninf.2011.00023

Zhang, B., Yang, G., Xu, C., Zhang, R., He, X., & Hu, W. (2024). The volume and structural covariance network of thalamic nuclei in patients with Wilson’s disease: An investigation of the association with neurological impairment. Neurological Sciences, 45(5), 2063–2073. 10.1007/s10072-023-07245-2

