## Supplementary Information for "Toward Reliable Thalamic Segmentation: a rigorous evaluation of automated methods for structural MRI"

### Supplementary Table 1

| **Dataset** | **Subject** | **Total thalamic volume (n vox)** | | |
| --- | --- | --- | --- | --- |
|  |  | **FS-DTI** | **FS-T1** | **HIPS-THOMAS** |
| HCP35 | mgh_1001 | 10753 | 12213 | 9845 |
| HCP35 | mgh_1002 | 11830 | 12818 | 10495 |
| HCP35 | mgh_1003 | 12958 | 14627 | 11598 |
| HCP35 | mgh_1004 | 13451 | 12453 | 11470 |
| HCP35 | mgh_1005 | 10551 | 11952 | 10063 |
| HCP35 | mgh_1006 | 13679 | 12926 | 11523 |
| HCP35 | mgh_1007 | 12453 | 16887 | 13117 |
| HCP35 | mgh_1008 | 13937 | 13333 | 11608 |
| HCP35 | mgh_1009 | 13456 | 13674 | 11892 |
| HCP35 | mgh_1010 | 13094 | 12317 | 10945 |
| HCP35 | mgh_1011 | 11961 | 12720 | 10850 |
| HCP35 | mgh_1012 | 13439 | 12630 | 10835 |
| HCP35 | mgh_1013 | 13073 | 13128 | 10906 |
| HCP35 | mgh_1014 | 15556 | 14239 | 11815 |
| HCP35 | mgh_1015 | 11973 | 13966 | 10926 |
| HCP35 | mgh_1016 | 14451 | 13179 | 11845 |
| HCP35 | mgh_1017 | 11372 | 11709 | 9745 |
| HCP35 | mgh_1018 | 15352 | 14261 | 12199 |
| HCP35 | mgh_1019 | 12411 | 13142 | 11566 |
| HCP35 | mgh_1020 | 2957 | 14504 | 11481 |
| HCP35 | mgh_1021 | 11739 | 11698 | 9825 |
| HCP35 | mgh_1022 | 14479 | 14551 | 13724 |
| HCP35 | mgh_1023 | 10332 | 11579 | 10104 |
| HCP35 | mgh_1024 | 11750 | 11920 | 10576 |
| HCP35 | mgh_1025 | 13467 | 14743 | 11742 |
| HCP35 | mgh_1026 | 12627 | 12487 | 11265 |
| HCP35 | mgh_1027 | 11716 | 14735 | 10817 |
| HCP35 | mgh_1028 | 10519 | 10779 | 8560 |
| HCP35 | mgh_1029 | 14338 | 16234 | 13018 |
| HCP35 | mgh_1030 | 12626 | 13268 | 11647 |
| HCP35 | mgh_1031 | 12228 | 15345 | 11887 |
| HCP35 | mgh_1032 | 10804 | 10185 | 9370 |
| HCP35 | mgh_1033 | 12473 | 12498 | 11285 |
| HCP35 | mgh_1034 | 15600 | 16408 | 13747 |
| HCP35 | mgh_1035 | n/a | n/a | n/a |
| MAP35 | C1 | 15611 | 14490 | 12833 |
| MAP35 | C2 | 14720 | 13574 | 12220 |
| MAP35 | C3 | 10807 | 12314 | 9900 |
| MAP35 | C4 | 11257 | 11782 | 9341 |
| MAP35 | C5 | 13516 | 14023 | 11092 |
| MAP35 | C6 | 10117 | 12078 | 8905 |
| MAP35 | C7 | 11986 | 12294 | 9665 |
| MAP35 | C8 | 14025 | 12616 | 11455 |
| MAP35 | C9 | 10186 | 11062 | 9175 |
| MAP35 | C10 | 11870 | 12240 | 9568 |
| MAP35 | C11 | 11504 | 13056 | 9933 |
| MAP35 | C12 | 13076 | 13087 | 10686 |
| MAP35 | C13 | 14485 | 14309 | 11702 |
| MAP35 | C14 | 15961 | 15396 | 12791 |
| MAP35 | C15 | 10829 | 11950 | 9931 |
| MAP35 | C16 | 11288 | 12052 | 9929 |
| MAP35 | C17 | 10627 | 11994 | 9614 |
| MAP35 | C18 | 10955 | 12277 | 9545 |
| MAP35 | C19 | 10953 | 11494 | 9045 |
| MAP35 | C20 | 11747 | 13213 | 10485 |
| MAP35 | C21 | 11105 | 13234 | 9779 |
| MAP35 | C22 | 13600 | 13646 | 11742 |
| MAP35 | C23 | 10618 | 11697 | 9264 |
| MAP35 | C24 | 11075 | 12348 | 9824 |
| MAP35 | C25 | 13763 | 14661 | 12251 |
| MAP35 | C26 | 13154 | 14967 | 12082 |
| MAP35 | C27 | 13348 | 14196 | 11572 |
| MAP35 | C28 | 13340 | 14358 | 11519 |
| MAP35 | C29 | 12838 | 12704 | 10412 |
| MAP35 | C30 | 14628 | 14575 | 12673 |
| MAP35 | C31 | 15121 | 14064 | 11956 |
| MAP35 | C32 | 9524 | 9732 | 8393 |
| MAP35 | C33 | 10954 | 11380 | 10142 |
| MAP35 | C34 | 14338 | 12768 | 11535 |
| MAP35 | C35 | 15459 | 14741 | 12612 |

Total thalamic volumes derived from the three segmentation methods. FS-DTI: FreeSurfer’s joint segmentation of thalamic nuclei from T1 scan and DTI (Tregidgo et al., 2023); HIPS-THOMAS: Thalamus Optimized Multi-atlas Segmentation using Histogram-based Polynomial Synthesis (Vidal et al., 2024); MAP35, HCP35: datasets with T1-weighted MRIs and diffusion MRI available

### Supplementary Table 2

| **HIPS-THOMAS** | **FS-T1** | **FS-DTI** | **K-M** |
| --- | --- | --- | --- |
| AV | AV | AV | AV |
| CL | CL | CL | CL |
| CM | CM | CM | CM |
| LGN | LGN | LGN | LGNmc+LGNpc |
| MD-Pf | MDl+ MDm + Pf | MDl+ MDm + Pf | Pf + sPf + MDmc + MDpc |
| MGN | MGN | MGN | MGN |
| Pul | PuA+PuI+PuL+PuM | PuA+PuI+PuL+PuMl+PuMm | PuA+PuI+PuL+PuM |
| VA | VAmc + VApc | VAmc + VApc | VAmc + VApc |
| VLa | VLa | VLa | VLa |
| VLp | VLp | VLp | VLpd + VLpv |
| VPL | VPL | VPL | VPLa+VPLp |

Method for combining FreeSurfer and K-M nuclei to match the Morel nomenclature used by THOMAS, producing a unified space for thalamic segmentation comparison; **key:** AV: anteroventral nucleus; VA: ventral anterior nucleus; VLa: Ventrolateral anterior nucleus; VLp: Ventrolateral posterior nucleus; MD-Pf: mediodorsal-parafascicular nuclei; Pul: pulvinar nucleus; VPL: Ventral Posterolateral nucleus; CL: Centrolateral nucleus; CM: Centromedian nucleus; LGN: Lateral Geniculate Nucleus; MGN: Medial Geniculate Nucleus; MDl: mediodorsal nucleus – lateral portion; MDm: mediodorsal nucleus – medial portion; MDmc: mediodorsal nucleus – magnocellular portion; MDpc: mediodorsal nucleus – parvocellular portion; Pf: parafascicular nucleus; PuA: anterior pulvinar; PuI: inferior pulvinar; PuL: lateral pulvinar; PuM: medial pulvinar; PuMl: medial pulvinar (lateral segment for FS-DTI); PuMm: medial pulvinar (medial segment for FS-DTI) VAmc: ventral anterior magnocellular; VApc: ventral anterior parvocellular; LGNmc: lateral geniculate magnocellular; LGNpc: lateral geniculate parvocellular; sPf: subparafascicular; VLpd: ventrolateral posterior nucleus (dorsal) ; VLpv: ventrolateral posterior nucleus (ventral); VPLa: ventral posterolateral nucleus (anterior); VPLp: ventral posterolateral nucleus (posterior); L/R: Left, Right hemisphere; FS-T1: T_1_-based FreeSurfer segmentation (Iglesias et al., 2018); FS-DTI: FreeSurfer’s joint segmentation of thalamic nuclei from T1 scan and DTI (Tregidgo et al., 2023); HIPS-THOMAS: Thalamus Optimized Multi-atlas Segmentation using Histogram-based Polynomial Synthesis (Vidal et al., 2024); MAP35, HCP35: datasets with T1-weighted MRIs and diffusion MRI available; K-M: Krauth-Morel atlas (Krauth et al., 2010)

### Supplementary Table 3

| **7TAMIbrainDGN** | **K-M** | **HIPS-THOMAS** | **FS-T1** | **FS-DTI** |
| --- | --- | --- | --- | --- |
| AV | AV + AM + AD + LD | AV | AV | AV |
| CL | CL | CL | CL | CL |
| CM | CM | CM | CM | CM |
| LGN | LGNmc + LGNpc | LGN | LGN | LGN |
| MD | MDmc + MDpc | MD-Pf | MDl+ MDm + Pf | MDl+ MDm + Pf |
| MGN | MGN | MGN | MGN | MGN |
| Pu+PuA | (PuM + PuI + PuL)+ PuA | Pul | PuA+PuI+PuL+PuM | PuA+PuI+PuL+PuMl+PuMm |
| LT | VPM+VPLa+VPLp+VLa+VLpd+VLpv+  VM+VAmc+VApc+VPI | VPL+VLa+VLp+VA | VPL+VLa+VLp+VAmc+VApc | VPL+VLa+VLp+VAmc+VApc |

Method for combining FreeSurfer and HIPS-THOMAS nuclei to match the segmentations of interest in the 7TAMIbrainDGN atlas (Brun et al., 2022); **key:** AD: anterodorsal nucleus; AM: anteromedial nucleus; AV: anterior segmentation (7TAMIbrainDGN atlas) or anteroventral nucleus (all other columns); CL: Centrolateral nucleus; CM: Centromedian nucleus; FS-DTI: FreeSurfer’s joint segmentation of thalamic nuclei from T1 scan and DTI (Tregidgo et al., 2023); FS-T1: T_1_-based FreeSurfer segmentation (Iglesias et al., 2018); HIPS-THOMAS: Thalamus Optimized Multi-atlas Segmentation using Histogram-based Polynomial Synthesis (Vidal et al., 2024); K-M: Krauth-Morel atlas (Krauth et al., 2010); L/R: Left, Right hemisphere; LD: laterodorsal nucleus; LGN: Lateral Geniculate Nucleus; LGNmc: lateral geniculate magnocellular nucleus; LGNpc: lateral geniculate parvocellular nucleus; LT: lateral nuclei (7TAMIbrainDGN atlas); MAP35, HCP35: datasets with T1-weighted MRIs and diffusion MRI available; MD-Pf: mediodorsal-parafascicular nuclei; MD: mediodorsal nucleus (7TAMIbrainDGN atlas); MDl: mediodorsal nucleus – lateral portion; MDm: mediodorsal nucleus – medial portion; MDmc: mediodorsal nucleus – magnocellular portion; MDpc: mediodorsal nucleus – parvocellular portion; MGN: Medial Geniculate Nucleus; Pf: parafascicular nucleus; Pu: Pulvinar (7TAMIbrainDGN atlas), including PuM, PuI, and PuL nucleus; PuA: anterior pulvinar nucleus; PuI: inferior pulvinar nucleus; PuL: lateral pulvinar nucleus; Pul: pulvinar nucleus; Pul: Pulvinar nucleus, in all other columns; PuM: medial pulvinar nucleus; PuMl: medial pulvinar nucleus (lateral segment for FS-DTI); PuMm: medial pulvinar nucleus (medial segment for FS-DTI); sPf: subparafascicular nucleus; VA: ventral anterior nucleus; VAmc: ventral anterior magnocellular; VApc: ventral anterior parvocellular; VLa: Ventrolateral anterior nucleus; VLp: Ventrolateral posterior nucleus; VLpd: ventrolateral posterior nucleus (dorsal); VLpv: ventrolateral posterior nucleus (ventral); VM: ventromedial VPL: Ventral Posterolateral nucleus; VPLa: ventral posterolateral nucleus (anterior); VPLp: ventral posterolateral nucleus (posterior); VPM: ventral posteromedial nucleus.

### Supplementary Table 4

| **Seg.** | **Hem.** | **HCP35** | | | | | | | | | **MAP35** | | | | | | | | |
| --- | --- | --- | --- | --- | --- | --- | --- | --- | --- | --- | --- | --- | --- | --- | --- | --- | --- | --- | --- |
|  |  | **F** | **p-corr(22)** | **ges** | **HIPS-THOMAS vs. FS-T1** | | **HIPS-THOMAS vs. FS-DTI** | | **FS-T1 vs. FS-DTI** | | **F** | **p-corr(22)** | **ges** | **HIPS-THOMAS vs. FS-T1** | | **HIPS-THOMAS vs. FS-DTI** | | **FS-T1 vs. FS-DTI** | |
|  |  |  |  |  | **t** | **p-corr(3)** | **t** | **p-corr(3)** | **t** | **p-corr(3)** |  |  |  | **t** | **p-corr(3)** | **t** | **p-corr(3)** | **t** | **p-corr(3)** |
| **AV** | **L** | 37.74 | 6.E-09 | 0.33 | 7.14 | < 0.001 | 0.06 | 0.953 | -6.51 | < 0.001 | 1.42 | 0.498 | 0.01 | 1.17 | 0.756 | 1.06 | 0.756 | -0.34 | 0.756 |
|  | **R** | 19.43 | 1.E-05 | 0.29 | 3.95 | 0.001 | 6.40 | < 0.001 | 2.55 | 0.016 | 52.26 | < 0.001 | 0.29 | 5.76 | < 0.001 | 7.51 | < 0.001 | 0.76 | 0.453 |
| **CL** | **L** | 807.60 | <1.E-12 | 0.92 | 29.64 | < 0.001 | 19.96 | < 0.001 | -8.95 | < 0.001 | 381.19 | < 0.001 | 0.86 | 18.81 | < 0.001 | 16.39 | < 0.001 | -2.12 | 0.042 |
|  | **R** | 383.88 | <1.E-12 | 0.85 | 19.95 | <0.001 | 13.67 | <0.001 | -5.38 | <0.001 | 331.44 | < 0.001 | 0.84 | 17.63 | < 0.001 | 17.62 | < 0.001 | -2.12 | 0.042 |
| **CM** | **L** | 70.26 | 2.E-12 | 0.45 | 3.24 | 0.003 | -5.21 | < 0.001 | -9.91 | < 0.001 | 7.30 | 0.004 | 0.09 | -0.72 | 0.478 | -3.18 | 0.009 | -2.50 | 0.034 |
|  | **R** | 42.41 | 6.E-09 | 0.28 | -5.04 | < 0.001 | -5.39 | < 0.001 | 0.20 | 0.840 | 13.59 | < 0.001 | 0.09 | -0.26 | 0.793 | -2.91 | 0.019 | -2.64 | 0.025 |
| **LGN** | **L** | 487.55 | <1.E-12 | 0.88 | 24.72 | < 0.001 | 22.64 | < 0.001 | -3.81 | 0.001 | 205.09 | < 0.001 | 0.77 | 12.68 | < 0.001 | 16.34 | < 0.001 | 6.24 | < 0.001 |
|  | **R** | 120.22 | <1.E-12 | 0.66 | 12.69 | < 0.001 | 12.60 | < 0.001 | 4.43 | < 0.001 | 210.22 | < 0.001 | 0.73 | 13.44 | < 0.001 | 17.83 | < 0.001 | 2.23 | 0.032 |
| **MD-Pf** | **L** | 201.23 | <1.E-12 | 0.72 | -5.02 | < 0.001 | -17.10 | < 0.001 | -9.56 | < 0.001 | 196.18 | < 0.001 | 0.72 | -7.34 | < 0.001 | -18.51 | < 0.001 | -8.78 | < 0.001 |
|  | **R** | 98.05 | <1.E-12 | 0.37 | -6.34 | < 0.001 | -6.40 | < 0.001 | -2.10 | 0.044 | 62.50 | < 0.001 | 0.40 | 1.49 | 0.144 | -7.67 | < 0.001 | -6.80 | < 0.001 |
| **MGN** | **L** | 92.57 | <1.E-12 | 0.68 | 2.33 | 0.026 | -19.29 | < 0.001 | -11.61 | < 0.001 | 69.58 | < 0.001 | 0.52 | -10.09 | < 0.001 | -12.17 | < 0.001 | 0.84 | 0.406 |
|  | **R** | 89.89 | 1.E-12 | 0.62 | -9.40 | < 0.001 | -15.91 | < 0.001 | -1.12 | 0.273 | 136.97 | < 0.001 | 0.61 | -6.36 | < 0.001 | -12.71 | < 0.001 | -5.68 | < 0.001 |
| **Pul** | **L** | 414.13 | <1.E-12 | 0.80 | 21.02 | < 0.001 | 10.14 | < 0.001 | -11.14 | < 0.001 | 268.52 | < 0.001 | 0.69 | 15.26 | < 0.001 | 9.64 | < 0.001 | -4.45 | < 0.001 |
|  | **R** | 71.54 | <1.E-12 | 0.32 | 6.60 | < 0.001 | 3.22 | 0.003 | -3.80 | 0.001 | 47.19 | < 0.001 | 0.20 | 4.71 | < 0.001 | 2.67 | 0.023 | -2.46 | 0.023 |
| **VA** | **L** | 21.71 | 6.E-06 | 0.27 | -0.54 | 6.E-01 | -7.59 | 4.E-08 | -4.42 | 2.E-04 | 3.87 | 0.129 | 0.06 | -2.12 | 0.082 | -2.88 | 0.020 | -0.28 | 0.780 |
|  | **R** | 13.39 | 3.E-05 | 0.10 | 3.83 | 1.E-03 | 4.85 | 9.E-05 | -0.17 | 9.E-01 | 65.14 | < 0.001 | 0.38 | 6.41 | < 0.001 | 7.72 | < 0.001 | 2.50 | 0.017 |
| **Vla** | **L** | 195.55 | <1.E-12 | 0.72 | 19.53 | < 0.001 | 7.86 | < 0.001 | -8.23 | < 0.001 | 33.44 | < 0.001 | 0.37 | 11.15 | < 0.001 | 5.09 | < 0.001 | -3.13 | 0.004 |
|  | **R** | 8.93 | 2.E-03 | 0.13 | 3.08 | 0.009 | 3.40 | 0.005 | 1.51 | 0.140 | 1.19 | 0.498 | 0.01 | -1.08 | 0.864 | -1.03 | 0.864 | -0.19 | 0.864 |
| **VLp** | **L** | 301.78 | <1.E-12 | 0.79 | 22.87 | < 0.001 | 11.63 | < 0.001 | -5.94 | < 0.001 | 132.44 | < 0.001 | 0.65 | 16.22 | < 0.001 | 13.74 | < 0.001 | -1.76 | 0.087 |
|  | **R** | 58.68 | 8.E-10 | 0.51 | 11.13 | < 0.001 | 9.03 | < 0.001 | 4.25 | < 0.001 | 103.80 | < 0.001 | 0.63 | 16.08 | < 0.001 | 13.17 | < 0.001 | -1.26 | 0.215 |
| **VPL** | **L** | 272.42 | <1.E-12 | 0.79 | 17.76 | < 0.001 | 5.50 | < 0.001 | -14.70 | < 0.001 | 111.12 | < 0.001 | 0.56 | 7.38 | < 0.001 | 11.29 | < 0.001 | 4.08 | < 0.001 |
|  | **R** | 16.93 | 2.E-05 | 0.15 | 3.49 | 0.004 | 3.14 | 0.007 | 0.51 | 0.614 | 219.98 | < 0.001 | 0.70 | 12.66 | < 0.001 | 14.94 | < 0.001 | 3.27 | 0.002 |

Comparison of the three segmentation methods on Dice coefficients (native space) for the HCP35 and MAP35 datasets, for nuclei segmented by all three methods, relative to the Krauth-Morel atlas (Krauth et al., 2010); **key:** purple: FS-T1 and FS-DTI are joint-best; orange: HIPS-THOMAS and FS-DTI are joint-best; yellow: HIPS-THOMAS and FS-T1 are joint-best; grey: no best or joint-best method; blue: FS-DTI is best; green: HIPS-THOMAS is best; AV: anteroventral nucleus; VA: ventral anterior nucleus; VLa: Ventrolateral anterior nucleus; VLp: Ventrolateral posterior nucleus; MD-Pf: mediodorsal-parafascicular nuclei; Pul: pulvinar nucleus; VPL: Ventral Posterolateral nucleus; CL: Centrolateral nucleus; CM: Centromedian nucleus; LGN: Lateral Geniculate Nucleus; MGN: Medial Geniculate Nucleus; L/R: Left, Right hemisphere; FS-T1: T_1_-based FreeSurfer segmentation (Iglesias et al., 2018); FS-DTI: FreeSurfer’s joint segmentation of thalamic nuclei from T1 scan and DTI (Tregidgo et al., 2023); HIPS-THOMAS: Thalamus Optimized Multi-atlas Segmentation using Histogram-based Polynomial Synthesis (Vidal et al., 2024); MAP35, HCP35: datasets with T1-weighted MRIs and diffusion MRI available; p-corr: the p-value was adjusted to correct for the number of multiple comparisons made (n=22), using the Holm-Bonferroni sequential correction method (Holm, 1979); ges: generalized eta squared.

### Supplementary Table 5

| **Seg.** | **Hem.** | **HCP35** | | | | | | | | | **MAP35** | | | | | | | | |
| --- | --- | --- | --- | --- | --- | --- | --- | --- | --- | --- | --- | --- | --- | --- | --- | --- | --- | --- | --- |
|  |  | **F** | **p-corr(16)** | **ges** | **HIPS-THOMAS vs. FS-T1** | | **HIPS-THOMAS vs. FS-DTI** | | **FS-T1 vs. FS-DTI** | | **F** | **p-corr(16)** | **ges** | **HIPS-THOMAS vs. FS-T1** | | **HIPS-THOMAS vs. FS-DTI** | | **FS-T1 vs. FS-DTI** | |
|  |  |  |  |  | **t(32)** | **p-corr(3)** | **t(32)** | **p-corr(3)** | **t(32)** | **p-corr(3)** |  |  |  | **t(34)** | **p-corr(3)** | **t(34)** | **p-corr(3)** | **t(34)** | **p-corr(3)** |
| **AV** | **L** | 167.71 | <1E-12 | 0.69 | 12.42 | 3.E-13 | 9.94 | 5.E-11 | -5.26 | 9.E-06 | 130.71 | <1E-12 | 0.45 | 9.25 | 2.E-10 | 16.62 | 2.E-17 | 6.05 | 7.E-07 |
|  | **R** | 74.47 | <1E-12 | 0.43 | 6.77 | 2.E-07 | 7.18 | 1.E-07 | -0.22 | 8.E-01 | 122.90 | <1E-12 | 0.37 | 10.89 | 3.E-12 | 18.75 | 5.E-19 | 0.12 | 9.E-01 |
| **CL** | **L** | 700.18 | <1E-12 | 0.93 | 28.68 | 6.E-24 | 20.43 | 1.E-19 | -8.04 | 4.E-09 | 415.00 | <1E-12 | 0.87 | 20.29 | 3.E-20 | 21.11 | 1.E-20 | -1.46 | 2.E-01 |
|  | **R** | 350.49 | <1E-12 | 0.85 | 20.51 | 2.E-19 | 12.63 | 1.E-13 | -6.57 | 2.E-07 | 333.76 | <1E-12 | 0.85 | 20.15 | 5.E-20 | 17.69 | 2.E-18 | -3.85 | 5.E-04 |
| **CM** | **L** | 245.25 | <1E-12 | 0.73 | 13.62 | 2.E-14 | 11.20 | 3.E-12 | -5.44 | 5.E-06 | 488.29 | <1E-12 | 0.82 | 23.96 | 1.E-22 | 26.18 | 1.E-23 | 2.51 | 2.E-02 |
|  | **R** | 26.72 | 8.E-06 | 0.30 | 3.44 | 2.E-03 | 6.01 | 3.E-06 | 4.39 | 2.E-04 | 214.82 | <1E-12 | 0.68 | 15.14 | 3.E-16 | 15.04 | 3.E-16 | 0.67 | 5.E-01 |
| **LGN** | **L** | 350.50 | <1E-12 | 0.86 | 21.62 | 3.E-20 | 20.52 | 1.E-19 | -1.98 | 6.E-02 | 445.58 | <1E-12 | 0.83 | 19.25 | 1.E-19 | 26.40 | 9.E-24 | 9.17 | 1.E-10 |
|  | **R** | 504.18 | <1E-12 | 0.84 | 20.75 | 1.E-19 | 17.53 | 1.E-17 | 2.92 | 6.E-03 | 575.07 | <1E-12 | 0.87 | 31.61 | 3.E-26 | 28.59 | 5.E-25 | 3.55 | 1.E-03 |
| **MD** | **L** | 156.72 | <1E-12 | 0.66 | 10.07 | 4.E-11 | -1.26 | 2.E-01 | -13.24 | 5.E-14 | 68.48 | <1E-12 | 0.45 | 10.53 | 9.E-12 | 3.84 | 5.E-04 | -10.03 | 2.E-11 |
|  | **R** | 39.54 | 1.E-09 | 0.27 | 5.10 | 3.E-05 | -0.63 | 5.E-01 | -6.47 | 9.E-07 | 118.67 | <1E-12 | 0.54 | 15.23 | 3.E-16 | 5.69 | 2.E-06 | -9.20 | 2.E-10 |
| **MGN** | **L** | 167.26 | <1E-12 | 0.77 | 12.24 | 3.E-13 | 24.93 | 5.E-22 | 2.94 | 6.E-03 | 83.65 | <1E-12 | 0.59 | 7.59 | 2.E-08 | 16.54 | 2.E-17 | 3.26 | 3.E-03 |
|  | **R** | 128.25 | <1E-12 | 0.68 | 12.78 | 8.E-14 | 17.83 | 1.E-17 | 0.50 | 6.E-01 | 109.17 | <1E-12 | 0.51 | 6.73 | 1.E-07 | 15.60 | 1.E-16 | 8.63 | 9.E-10 |
| **Pul** | **L** | 383.86 | <1E-12 | 0.79 | 20.90 | 1.E-19 | 11.95 | 5.E-13 | -9.75 | 4.E-11 | 327.41 | <1E-12 | 0.72 | 24.04 | 2.E-22 | 19.34 | 1.E-19 | -4.26 | 2.E-04 |
|  | **R** | 292.04 | <1E-12 | 0.69 | 15.25 | 9.E-16 | 9.39 | 2.E-10 | -4.18 | 2.E-04 | 193.55 | <1E-12 | 0.64 | 15.47 | 1.E-16 | 17.23 | 7.E-18 | -3.92 | 4.E-04 |
| **LT** | **L** | 231.78 | <1E-12 | 0.75 | 14.93 | 2.E-15 | 1.38 | 2.E-01 | -11.61 | 1.E-12 | 55.39 | 2.E-11 | 0.43 | 12.93 | 3.E-14 | 9.50 | 8.E-11 | 0.14 | 9.E-01 |
|  | **R** | 11.17 | 1.E-03 | 0.10 | -0.65 | 5.E-01 | 2.15 | 8.E-02 | 3.05 | 1.E-02 | 74.73 | <1E-12 | 0.40 | 13.64 | 7.E-15 | 10.83 | 3.E-12 | 1.73 | 9.E-02 |

Comparison of the three segmentation methods on Dice coefficients (native space) for the HCP35 and MAP35 datasets, for nuclei segmented by all three methods, relative to 7TAMI; **key:** orange: HIPS-THOMAS and FS-DTI are joint-best; grey: no best or joint-best method; green: HIPS-THOMAS is best; AV: “anteroventral” nucleus; LT: lateral thalamic nuclei; MD: “mediodorsal” nucleus; Pul: pulvinar nucleus; CL: Centrolateral nucleus; CM: Centromedian nucleus; LGN: Lateral Geniculate Nucleus; MGN: Medial Geniculate Nucleus; L/R: Left, Right hemisphere; FS-T1: T_1_-based FreeSurfer segmentation (Iglesias et al., 2018); FS-DTI: FreeSurfer’s joint segmentation of thalamic nuclei from T1 scan and DTI (Tregidgo et al., 2023); HIPS-THOMAS: Thalamus Optimized Multi-atlas Segmentation using Histogram-based Polynomial Synthesis (Vidal et al., 2024); MAP35, HCP35: datasets with T1-weighted MRIs and diffusion MRI available; p-corr: the p-value was adjusted to correct for the number of multiple comparisons made (n=16), using the Holm-Bonferroni sequential correction method (Holm, 1979); ges: generalized eta squared; 7TAMI: the custom-made MNI version of the atlas of deep grey matter nuclei, part of the 7TAMIbrain dataset (Brun et al., 2022).

### Supplementary Table 6

| **Seg.** | **Hem.** | HCP35 | | | | | | | | | MAP35 | | | | | | | | |
| --- | --- | --- | --- | --- | --- | --- | --- | --- | --- | --- | --- | --- | --- | --- | --- | --- | --- | --- | --- |
|  |  | ANOVA | | | HIPS-THOMAS vs. FS-T1 | | HIPS-THOMAS vs. FS-DTI | | FS-T1 vs. FS-DTI | | ANOVA | | | HIPS-THOMAS vs. FS-T1 | | HIPS-THOMAS vs. FS-DTI | | FS-T1 vs. FS-DTI | |
|  |  | **F** | **ges** | **p-corr(22)** | **t(32)** | **p-corr(3)** | **t(32)** | **p-corr(3)** | **t(32)** | **p-corr(3)** | **F** | **ges** | **p-corr(22)** | **t(34)** | **p-corr(3)** | **t(34)** | **p-corr(3)** | **t(34)** | **p-corr(3)** |
| AV | L | 5.92 | 0.10 | 1.E-02 | -3.41 | 0.005 | -2.69 | 0.023 | 0.14 | 0.888 | 11.35 | 0.08 | <0.001 | 0.17 | 0.868 | -2.92 | 0.019 | -2.85 | 0.019 |
|  | R | 25.33 | 0.34 | 6.E-08 | -2.65 | 0.012 | -8.72 | <0.001 | -4.15 | <0.001 | 47.76 | 0.22 | <0.001 | -4.26 | <0.001 | -5.83 | <0.001 | -1.80 | 0.082 |
| CL | L | 377.33 | 0.86 | <1.E-12 | -17.94 | <0.001 | -43.17 | <0.001 | 0.93 | 0.359 | 38.28 | 0.43 | <0.001 | -4.08 | <0.001 | -7.07 | <0.001 | -16.46 | <0.001 |
|  | R | 323.80 | 0.83 | <1.E-12 | -16.89 | <0.001 | -28.61 | <0.001 | 0.89 | 0.383 | 45.58 | 0.47 | <0.001 | -4.65 | <0.001 | -7.78 | <0.001 | -10.65 | <0.001 |
| CM | L | 51.05 | 0.38 | 4.E-09 | -1.91 | 0.065 | 5.87 | <0.001 | 7.87 | <0.001 | 15.73 | 0.18 | <0.001 | 2.31 | 0.054 | 5.36 | <0.001 | 2.07 | 0.054 |
|  | R | 41.99 | 0.33 | 3.E-08 | 5.61 | <0.001 | 6.84 | <0.001 | 0.71 | 0.481 | 15.15 | 0.11 | <0.001 | 1.49 | 0.146 | 3.88 | 0.001 | 2.00 | 0.107 |
| LGN | L | 448.46 | 0.88 | <1.E-12 | -25.56 | <0.001 | -23.09 | <0.001 | 8.61 | <0.001 | 252.82 | 0.82 | <0.001 | -21.89 | <0.001 | -20.43 | <0.001 | -2.85 | 0.007 |
|  | R | 80.63 | 0.57 | <1.E-12 | -14.82 | <0.001 | -10.65 | <0.001 | -0.28 | 0.779 | 167.99 | 0.72 | <0.001 | -17.17 | <0.001 | -15.96 | <0.001 | -0.39 | 0.697 |
| MD-Pf | L | 272.35 | 0.78 | <1.E-12 | 9.40 | <0.001 | 20.36 | <0.001 | 7.95 | <0.001 | 199.22 | 0.79 | <0.001 | 9.97 | <0.001 | 20.26 | <0.001 | 9.30 | <0.001 |
|  | R | 116.32 | 0.46 | <1.E-12 | 5.42 | <0.001 | 8.59 | <0.001 | 4.19 | <0.001 | 66.56 | 0.46 | <0.001 | 2.01 | 0.053 | 10.61 | <0.001 | 6.05 | <0.001 |
| MGN | L | 43.29 | 0.47 | 6.E-08 | -3.12 | 0.004 | 9.19 | <0.001 | 8.26 | <0.001 | 80.13 | 0.51 | <0.001 | 9.80 | <0.001 | 6.92 | <0.001 | -3.45 | 0.002 |
|  | R | 88.42 | 0.61 | <1.E-12 | 10.73 | <0.001 | 9.55 | <0.001 | -2.56 | 0.015 | 100.22 | 0.55 | <0.001 | 6.66 | <0.001 | 9.99 | <0.001 | 2.51 | 0.017 |
| Pul | L | 382.62 | 0.82 | <1.E-12 | -19.92 | <0.001 | -7.02 | <0.001 | 13.65 | <0.001 | 193.55 | 0.57 | <0.001 | -11.74 | <0.001 | -9.31 | <0.001 | 2.56 | 0.015 |
|  | R | 71.18 | 0.21 | <1.E-12 | -6.17 | <0.001 | -3.47 | 0.003 | 1.79 | 0.083 | 105.91 | 0.33 | <0.001 | -6.53 | <0.001 | -5.85 | <0.001 | 1.29 | 0.204 |
| VA | L | 20.58 | 0.21 | 2.E-05 | -0.56 | 6.E-01 | 8.31 | 5.E-09 | 4.72 | 9.E-05 | 5.45 | 0.07 | 0.019 | 3.75 | 0.002 | 0.43 | 0.667 | -1.78 | 0.168 |
|  | R | 7.10 | 0.06 | 6.E-03 | -2.42 | 4.E-02 | -3.16 | 1.E-02 | -1.68 | 1.E-01 | 45.35 | 0.26 | <0.001 | -3.81 | 0.001 | -5.65 | <0.001 | -2.71 | 0.010 |
| Vla | L | 182.95 | 0.70 | <1.E-12 | -15.26 | <0.001 | -6.67 | <0.001 | 8.78 | <0.001 | 63.14 | 0.53 | <0.001 | -12.58 | <0.001 | -1.93 | 0.063 | 8.80 | <0.001 |
|  | R | 11.64 | 0.18 | 1.E-03 | -5.29 | <0.001 | -3.50 | 0.003 | -0.04 | 0.970 | 19.91 | 0.15 | <0.001 | -4.15 | 0.001 | 0.39 | 0.702 | 3.49 | 0.003 |
| VLp | L | 139.33 | 0.68 | <1.E-12 | -13.32 | <0.001 | -13.58 | <0.001 | 3.18 | 0.003 | 74.64 | 0.57 | <0.001 | -10.87 | <0.001 | -11.98 | <0.001 | -1.65 | 0.107 |
|  | R | 70.68 | 0.58 | 9.E-10 | -4.87 | <0.001 | -9.40 | <0.001 | -7.65 | <0.001 | 126.12 | 0.60 | <0.001 | -11.45 | <0.001 | -14.26 | <0.001 | -3.44 | 0.002 |
| VPL | L | 311.17 | 0.81 | <1.E-12 | -16.78 | <0.001 | -3.50 | 0.001 | 15.22 | <0.001 | 115.08 | 0.66 | <0.001 | -13.56 | <0.001 | -12.04 | <0.001 | -3.00 | 0.005 |
|  | R | 17.68 | 0.18 | 2.E-05 | -5.36 | <0.001 | -2.68 | 0.023 | 1.12 | 0.270 | 163.60 | 0.69 | <0.001 | -14.43 | <0.001 | -13.99 | <0.001 | -3.05 | 0.004 |

Comparison of the three segmentation methods on AHDs (native space) for the HCP35 and MAP35 datasets for nuclei segmented by all three methods, relative to the Krauth-Morel atlas (Krauth et al., 2010). **key:** purple: FS-T1 and FS-DTI are joint-best; orange: HIPS-THOMAS and FS-DTI are joint-best; yellow: HIPS-THOMAS and FS-T1 are joint-best; grey: no best or joint-best method; blue: FS-DTI is best; green: HIPS-THOMAS is best; red: FS-T1 is best; AHD: average Hausdorff distance; AV: anteroventral nucleus; VA: ventral anterior nucleus; VLa: Ventrolateral anterior nucleus; VLp: Ventrolateral posterior nucleus; MD-Pf: mediodorsal-parafascicular nuclei; Pul: pulvinar nucleus; VPL: Ventral Posterolateral nucleus; CL: Centrolateral nucleus; CM: Centromedian nucleus; LGN: Lateral Geniculate Nucleus; MGN: Medial Geniculate Nucleus; L/R: Left, Right hemisphere; FS-T1: T_1_-based FreeSurfer segmentation (Iglesias et al., 2018); FS-DTI: FreeSurfer’s joint segmentation of thalamic nuclei from T1 scan and DTI (Tregidgo et al., 2023); HIPS-THOMAS: Thalamus Optimized Multi-atlas Segmentation using Histogram-based Polynomial Synthesis (Vidal et al., 2024); MAP35, HCP35: datasets with T1-weighted MRIs and diffusion MRI available. p-corr: the p-value was adjusted to correct for the number of multiple comparisons made (n=22), using the Holm-Bonferroni sequential correction method (Holm, 1979); ges: generalized eta squared.

### Supplementary Table 7

| **Seg.** | **Hem.** | **HCP35** | | | | | | | | | **MAP35** | | | | | | | | |
| --- | --- | --- | --- | --- | --- | --- | --- | --- | --- | --- | --- | --- | --- | --- | --- | --- | --- | --- | --- |
|  |  |  | **p-corr(16)** | **ges** | **HIPS-THOMAS vs. FS-T1** | | **HIPS-THOMAS vs. FS-DTI** | | **FS-T1 vs. FS-DTI** | |  | **p-corr(16)** | **ges** | **HIPS-THOMAS vs. FS-T1** | | **HIPS-THOMAS vs. FS-DTI** | | **FS-T1 vs. FS-DTI** | |
|  |  | **F** |  |  | **t(32)** | **p-corr(3)** | **t(32)** | **p-corr(3)** | **t(32)** | **p-corr(3)** | **F** |  |  | **t(34)** | **p-corr(3)** | **t(34)** | **p-corr(3)** | **t(34)** | **p-corr(3)** |
| **AV** | **L** | 135.45 | <1E-12 | 0.44 | -7.49 | 5.E-08 | -2.94 | 6.E-03 | 5.80 | 4.E-06 | 73.73 | <1E-12 | 0.33 | -8.25 | 3.E-09 | -10.82 | 4.E-12 | -5.12 | 1.E-05 |
|  | **R** | 72.47 | <1E-12 | 0.32 | -5.93 | 4.E-06 | -1.96 | 6.E-02 | 4.52 | 2.E-04 | 70.34 | <1E-12 | 0.25 | -10.94 | 2.E-12 | -11.13 | 2.E-12 | -1.32 | 2.E-01 |
| **CL** | **L** | 535.71 | <1E-12 | 0.89 | -20.73 | 8.E-20 | -53.23 | 2.E-32 | 0.44 | 7.E-01 | 52.08 | 2.E-08 | 0.50 | -5.16 | 1.E-05 | -8.48 | 1.E-09 | -16.57 | 2.E-17 |
|  | **R** | 429.63 | <1E-12 | 0.85 | -17.40 | 3.E-17 | -29.13 | 1.E-23 | 0.01 | 1.E+00 | 50.89 | 2.E-08 | 0.49 | -5.70 | 2.E-06 | -8.13 | 4.E-09 | -12.99 | 3.E-14 |
| **CM** | **L** | 220.05 | <1E-12 | 0.79 | -17.07 | 4.E-17 | -14.91 | 1.E-15 | 6.83 | 1.E-07 | 312.87 | <1E-12 | 0.84 | -18.98 | 2.E-19 | -25.43 | 3.E-23 | 1.74 | 9.E-02 |
|  | **R** | 45.95 | 7.E-10 | 0.47 | -8.00 | 8.E-09 | -8.76 | 2.E-09 | -0.50 | 6.E-01 | 258.83 | <1E-12 | 0.74 | -15.86 | 6.E-17 | -20.07 | 6.E-20 | 1.56 | 1.E-01 |
| **LGN** | **L** | 531.30 | <1E-12 | 0.89 | -27.07 | 3.E-23 | -27.82 | 2.E-23 | 4.96 | 2.E-05 | 697.12 | <1E-12 | 0.92 | -26.01 | 1.E-23 | -43.12 | 9.E-31 | -4.86 | 3.E-05 |
|  | **R** | 442.56 | <1E-12 | 0.85 | -24.98 | 4.E-22 | -21.97 | 1.E-20 | -0.19 | 9.E-01 | 578.81 | <1E-12 | 0.90 | -30.65 | 5.E-26 | -31.16 | 4.E-26 | -1.51 | 1.E-01 |
| **MD** | **L** | 217.76 | <1E-12 | 0.79 | -16.99 | 4.E-17 | -4.99 | 2.E-05 | 14.87 | 1.E-15 | 200.62 | <1E-12 | 0.78 | -17.96 | 1.E-18 | -6.02 | 8.E-07 | 19.15 | 3.E-19 |
|  | **R** | 121.70 | <1E-12 | 0.63 | -11.01 | 6.E-12 | -2.94 | 6.E-03 | 9.91 | 6.E-11 | 199.92 | <1E-12 | 0.75 | -17.02 | 1.E-17 | -6.30 | 4.E-07 | 15.46 | 1.E-16 |
| **MGN** | **L** | 411.16 | <1E-12 | 0.88 | -15.02 | 1.E-15 | -37.73 | 1.E-27 | -9.91 | 3.E-11 | 234.07 | <1E-12 | 0.81 | -9.11 | 1.E-10 | -20.86 | 2.E-20 | -12.90 | 2.E-14 |
|  | **R** | 255.25 | <1E-12 | 0.77 | -9.37 | 2.E-10 | -24.91 | 5.E-22 | -6.90 | 8.E-08 | 242.88 | <1E-12 | 0.77 | -10.82 | 1.E-12 | -18.95 | 4.E-19 | -12.95 | 2.E-14 |
| **Pul** | **L** | 190.42 | <1E-12 | 0.66 | -14.31 | 6.E-15 | -7.40 | 2.E-08 | 7.89 | 1.E-08 | 206.49 | <1E-12 | 0.51 | -16.58 | 2.E-17 | -15.09 | 2.E-16 | 2.58 | 1.E-02 |
|  | **R** | 174.65 | <1E-12 | 0.48 | -11.12 | 5.E-12 | -6.68 | 3.E-07 | 2.73 | 1.E-02 | 245.59 | <1E-12 | 0.56 | -18.40 | 9.E-19 | -16.20 | 3.E-17 | 5.45 | 4.E-06 |
| **LT** | **L** | 326.50 | <1E-12 | 0.79 | -16.25 | 2.E-16 | -3.25 | 3.E-03 | 13.10 | 4.E-14 | 164.39 | <1E-12 | 0.70 | -20.44 | 3.E-20 | -12.47 | 6.E-14 | 4.38 | 1.E-04 |
|  | **R** | 15.96 | 7.E-05 | 0.13 | -5.34 | 2.E-05 | -2.75 | 2.E-02 | 0.21 | 8.E-01 | 141.11 | <1E-12 | 0.55 | -19.72 | 1.E-19 | -10.09 | 2.E-11 | 4.44 | 9.E-05 |

Comparison of the three segmentation methods on AHDs (native space) for the HCP35 and MAP35 datasets, for nuclei segmented by all three methods, relative to 7TAMI; **key:** orange: HIPS-THOMAS and FS-DTI are joint-best; grey: no best or joint-best method; green: HIPS-THOMAS is best; AHD: average Hausdorff distance; AV: “anteroventral” nucleus; LT: lateral thalamic nuclei; MD: “mediodorsal” nucleus; Pul: pulvinar nucleus; CL: Centrolateral nucleus; CM: Centromedian nucleus; LGN: Lateral Geniculate Nucleus; MGN: Medial Geniculate Nucleus; L/R: Left, Right hemisphere; FS-T1: T_1_-based FreeSurfer segmentation (Iglesias et al., 2018); FS-DTI: FreeSurfer’s joint segmentation of thalamic nuclei from T1 scan and DTI (Tregidgo et al., 2023); HIPS-THOMAS: Thalamus Optimized Multi-atlas Segmentation using Histogram-based Polynomial Synthesis (Vidal et al., 2024); MAP35, HCP35: datasets with T1-weighted MRIs and diffusion MRI available; p-corr: the p-value was adjusted to correct for the number of multiple comparisons made (n=16), using the Holm-Bonferroni sequential correction method (Holm, 1979); ges: generalized eta squared; 7TAMI: the custom-made MNI version of the atlas of deep grey matter nuclei, part of the 7TAMIbrain dataset (Brun et al., 2022).

### Supplementary Table 8

| **Method** | **Ground Truth** | **Native space** | | | | **MNI space** | | | |
| --- | --- | --- | --- | --- | --- | --- | --- | --- | --- |
|  |  | **Dice≥0.6** | | **AHD≤1.0** | | **Dice≥0.6** | | **AHD≤1.0** | |
|  |  | **HCP35** | **MAP35** | **HCP35** | **MAP35** | **HCP35** | **MAP35** | **HCP35** | **MAP35** |
| **FS-DTI** | **K-M (/22)** | 5 | 3 | 8 | 5 | 8 | 7 | 9 | 7 |
|  | **7TAMI (/16)** | 5 | 6 | 6 | 5 | 6 | 6 | 6 | 4 |
| **FS-T1** | **K-M (/22)** | 2 | 2 | 8 | 8 | 7 | 6 | 8 | 10 |
|  | **7TAMI (/16)** | 4 | 4 | 4 | 6 | 4 | 6 | 3 | 3 |
| **HIPS-THOMAS** | **K-M (/22)** | 7 | 8 | 11 | 13 | 10 | 13 | 15 | 15 |
|  | **7TAMI (/16)** | 8 | 7 | 11 | 12 | 10 | 10 | 12 | 11 |

Number of segmentations per space (native/MNI), segmentation method (FS-T1/FS-DTI/HIPS-THOMAS), dataset (HCP35/MAP35) that have a Dice coefficient of ≥0.6 (at least “substantial” agreement/overlap) and/or a AHD≤1 (sufficiently small distance) relative to the corresponding K-M/7TAMI reference/ground-truth segmentation; **key:** FS-T1: T_1_-based FreeSurfer segmentation (Iglesias et al., 2018); FS-DTI: FreeSurfer’s joint segmentation of thalamic nuclei from T1 scan and DTI (Tregidgo et al., 2023); HIPS-THOMAS: Thalamus Optimized Multi-atlas Segmentation using Histogram-based Polynomial Synthesis (Vidal et al., 2024); MAP35, HCP35: datasets with T1-weighted MRIs and diffusion MRI available; AHD: average Hausdorff distance; K-M: Krauth-Morel atlas (Krauth et al., 2010); 7TAMI: the custom-made MNI version of the atlas of deep grey matter nuclei, part of the 7TAMIbrain dataset (Brun et al., 2022).

### Supplementary Figure 1

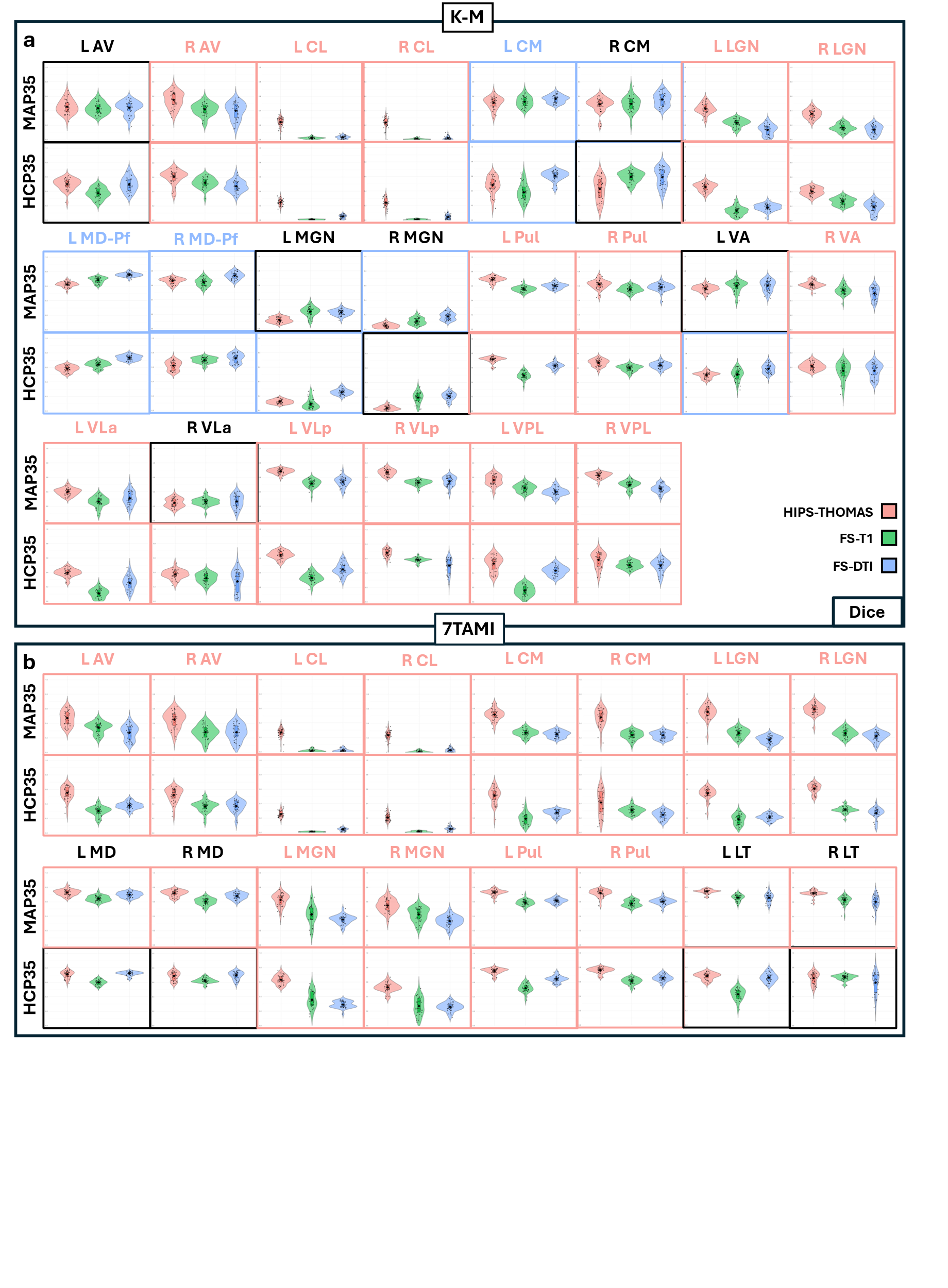

Violin plots of Dice overlap between nuclei segmented with FS-T1, FS-DTI, and HIPS-THOMAS relative to a) K-M and b) 7TAMI ground-truth reference segmentations (native space). Comparisons among methods that disclose significantly higher Dice coefficients for one method relative to the other two are indicated by correspondingly coloured panels; y-axis:0-1; **key:** AV: anteroventral nucleus; LT: lateral nuclei (7TAMI); VA: ventral anterior nucleus; VLa: Ventrolateral anterior nucleus; VLp: Ventrolateral posterior nucleus; MD-Pf: mediodorsal-parafascicular nuclei; Pul: pulvinar nucleus; VPL: Ventral Posterolateral nucleus; CL: Centrolateral nucleus; CM: Centromedian nucleus; LGN: Lateral Geniculate Nucleus; MGN: Medial Geniculate Nucleus; L/R: Left, Right hemisphere; FS-T1: T_1_-based FreeSurfer segmentation (Iglesias et al., 2018); FS-DTI: FreeSurfer’s joint segmentation of thalamic nuclei from T1 scan and DTI (Tregidgo et al., 2023); HIPS-THOMAS: Thalamus Optimized Multi-atlas Segmentation using Histogram-based Polynomial Synthesis (Vidal et al., 2024); MAP35, HCP35: datasets with T1w-MRIs and diffusion MRI available; black diamond: mean; black horizontal line: median; K-M: Krauth-Morel atlas (Krauth et al., 2010); 7TAMI: the custom-made MNI version of the atlas of deep grey matter nuclei, part of the 7TAMIbrain dataset (Brun et al., 2022).

### Supplementary Figure 2

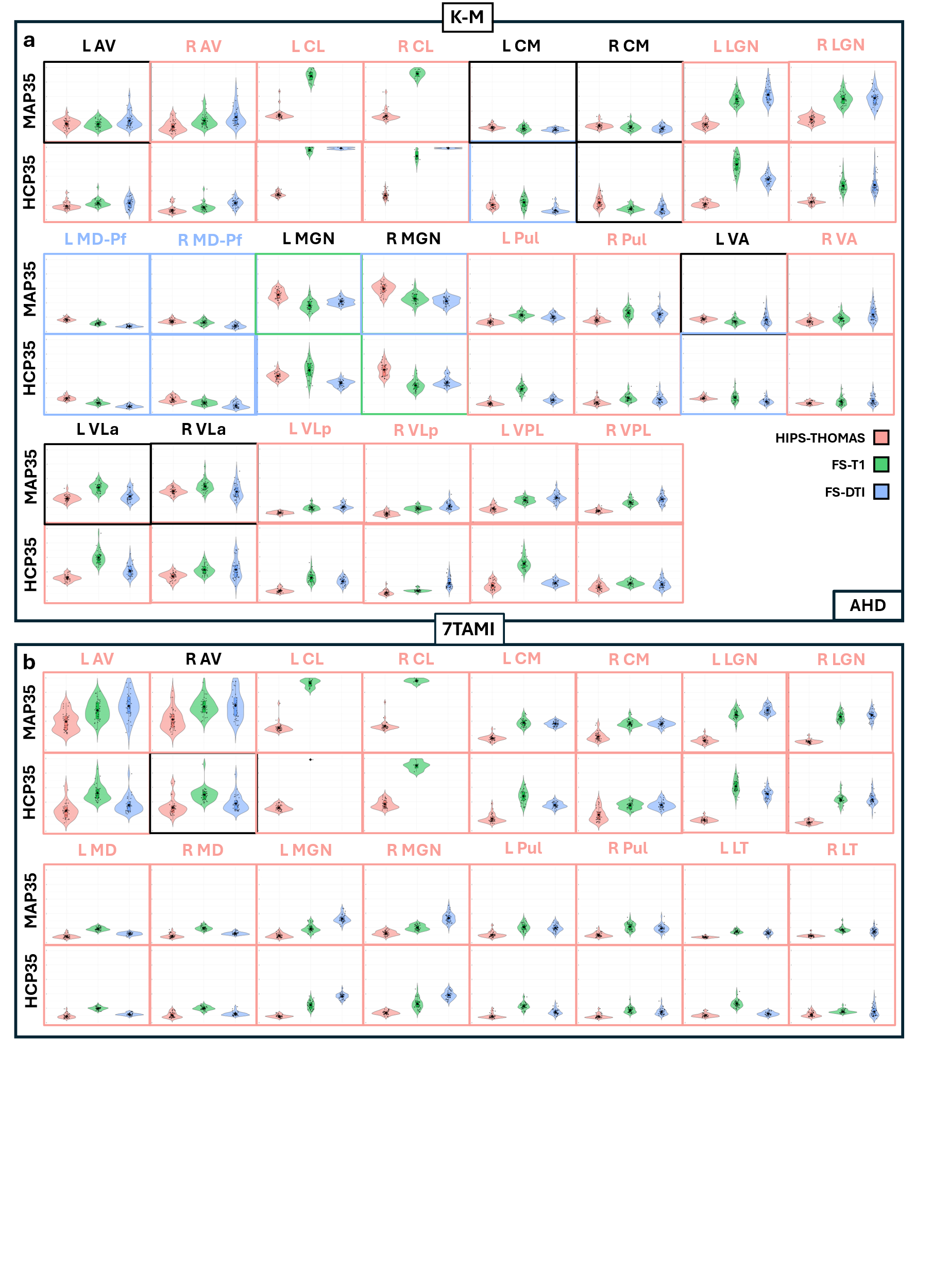

Violin plots of AHDs for native-space segmentations based on FS-T1, FS-DTI, and HIPS-THOMAS relative to a) K-M and b) 7TAMI ground-truth reference segmentations (y-axis:0-5mm, for display purposes). Comparisons among methods that disclose significantly lower AHD for one method relative to the other two are indicated by correspondingly coloured panels; **key:** AV: anteroventral nucleus; LT: lateral nuclei (7TAMI); VA: ventral anterior nucleus; VLa: Ventrolateral anterior nucleus; VLp: Ventrolateral posterior nucleus; MD-Pf: mediodorsal-parafascicular nuclei; Pul: pulvinar nucleus; VPL: Ventral Posterolateral nucleus; CL: Centrolateral nucleus; CM: Centromedian nucleus; LGN: Lateral Geniculate Nucleus; MGN: Medial Geniculate Nucleus; L/R: Left, Right hemisphere; FS-T1: T_1_-based FreeSurfer segmentation (Iglesias et al., 2018); FS-DTI: FreeSurfer’s joint segmentation of thalamic nuclei from T1 scan and DTI (Tregidgo et al., 2023); HIPS-THOMAS: Thalamus Optimized Multi-atlas Segmentation using Histogram-based Polynomial Synthesis (Vidal et al., 2024); AHD: Average Hausdorff Distance; MAP35, HCP35: datasets with T1-weighted MRIs and diffusion MRI available; black diamond: mean; black horizontal line: median. K-M: Krauth-Morel atlas (Krauth et al., 2010); 7TAMI: the custom-made MNI version of the atlas of deep grey matter nuclei, part of the 7TAMIbrain dataset (Brun et al., 2022).
